# Germline genetic risk converges through intercellular crosstalk in tumor microenvironment

**DOI:** 10.1101/2025.10.14.25337882

**Authors:** Xiaomeng Bai, Yang Liu, Shuai He, Chun-Ling Luo, Guo-Wang Lin, Dong-Mei Chi, Shuang-Yan Ye, Xiu-Zhi Wang, Yue-Feng Wen, You Zhou, Shu-Qiang Liu, Pan-Pan Wei, Xin-Yuan Guan, Hai-Qiang Mai, Soon Thye Lim, Jian-Jun Liu, Chiea Chuen Khor, Melvin Lee Kiang CHUA, Jin-Xin Bei, Yanni Zeng

## Abstract

Germline genetic variation contributes to nasopharyngeal carcinoma (NPC) risk, but how susceptibility genes act across tumor microenvironment (TME) and whether enriched cell types interact remain unclear. Here we integrated large-scale NPC genetic association data(N=9,447) with transcriptomic datasets of NPC tumors, including single-cell RNA-seq(N=31), bulk RNA-seq(N=180) and spatial transcriptomics(N=7). We found that common variant-derived risk was enriched in antigen-presenting cells, whereas rare variant-derived risk localized to epithelial and stromal populations. Strikingly, these genetically enriched cell types exhibited stronger intercellular interactions and closer spatial proximity than others, with the most prominent crosstalk observed between Langerhans dendritic cells(DC_C4_CD207, enriched for common variant risk) and HILPDA⁺ epithelial cells(EP_C10_HILPDA, enriched for rare variant risk). Clinically, combined infiltration of these two subtypes substantially improved prediction of patient prognosis. Together, our findings provide critical evidence that germline susceptibility shapes the TME in NPC and highlight intercellular crosstalk as a converging mechanism linking genetic risk to cancer pathogenesis.

## Introduction

Tumorigenesis is a multifactorial process shaped by the interplay of genetic factors, environmental exposures, and cellular components within the tumor microenvironment (TME), a critical niche for cancer initiation and progression^1,2^. Understanding how distinct cellular constituents operate within the TME, how malignant and non-malignant cells interact, and how germline genetic factors influence these dynamics is essential to deciphering cancer biology. In nasopharyngeal carcinoma (NPC), single-cell RNA sequencing (scRNA-seq) studies have revealed that in addition to malignant epithelial cells, the TME is characterized by extensive infiltration of immune cells (e.g., T cells, B cells, and myeloid cells) as well as stromal populations^3,4^. These diverse cell types engage in complex interactions that can either promote tumor progression or enhance anti-tumor responses^4^. Yet, how germline genetic factors contribute to, or underlie, these TME dynamics remains poorly understood.

Genetic predisposition is a well-established component of cancer etiology. Genome- and exome-wide association studies (GWAS and EWAS) have identified numerous cancer-related risk genes through common and rare variants^5,6^. In NPC, multiple susceptibility loci have been discovered across the allele frequency spectrum^7–9^. While coding germline mutations may exert direct effect on cells with malignant potential (epithelial cells in NPC), and thus have historically been the focus functional validation^10,11^, most cancer-associated variants map to non-coding regions^12^. This raises the possibility that germline variants modulate TME dynamics indirectly, through immune and stromal components in addition to malignant cells^12^. To fully capture these effects, it is critical to evaluate germline risk genes across malignant and malignant populations, and more importantly, to investigate how genetically influenced cell types interact within the TME. By far, the cellular contexts and interaction networks through which germline susceptibility shapes cancer risk remain largely unresolved^1,13^.

Recent advances in single-cell genetics have enabled systematic mapping of the cellular contexts through which genetic variants confer cancer risk^9,14,15^. By integrating genotype data with single cell transcriptomes, studies have identified cell types preferentially expressing GWAS-implicated genes, thereby refining the interpretation of cancer susceptibility loci^14,15^. Nonetheless, most single-cell genetic analyses focus on cell-intrinsic effects, overlooking the central role of intercellular interactions in the TME^14,16^. From the genetic perspective, interactions between TME components shaped by multiple germline variants represent a key intercellular context where polygenic risk converges. Yet, whether and how germline variants influence intercellular communication within the TME, particularly NPC, remains unknown.

Here, we address this gap by systematically investigating the intercellular communications among TME components enriched for common and rare genetic associations, using NPC as a model. By integrating large-scale NPC genetic association data with scRNA-seq, spatial transcriptomics, and bulk RNA-seq from tumor tissues, we uncover significant crosstalk between genetically enriched cell populations (**Fig. 1**). Importantly, we demonstrate that incorporating genetically influenced TME interactions improves the prediction of clinical prognosis. Our analyses establish a new paradigm in which germline variation shapes intercellular communication within the TME, providing novel insights into how genetic susceptibility loci contribute to cancer and complex disease at the intercellular level.

**Figure 1.**
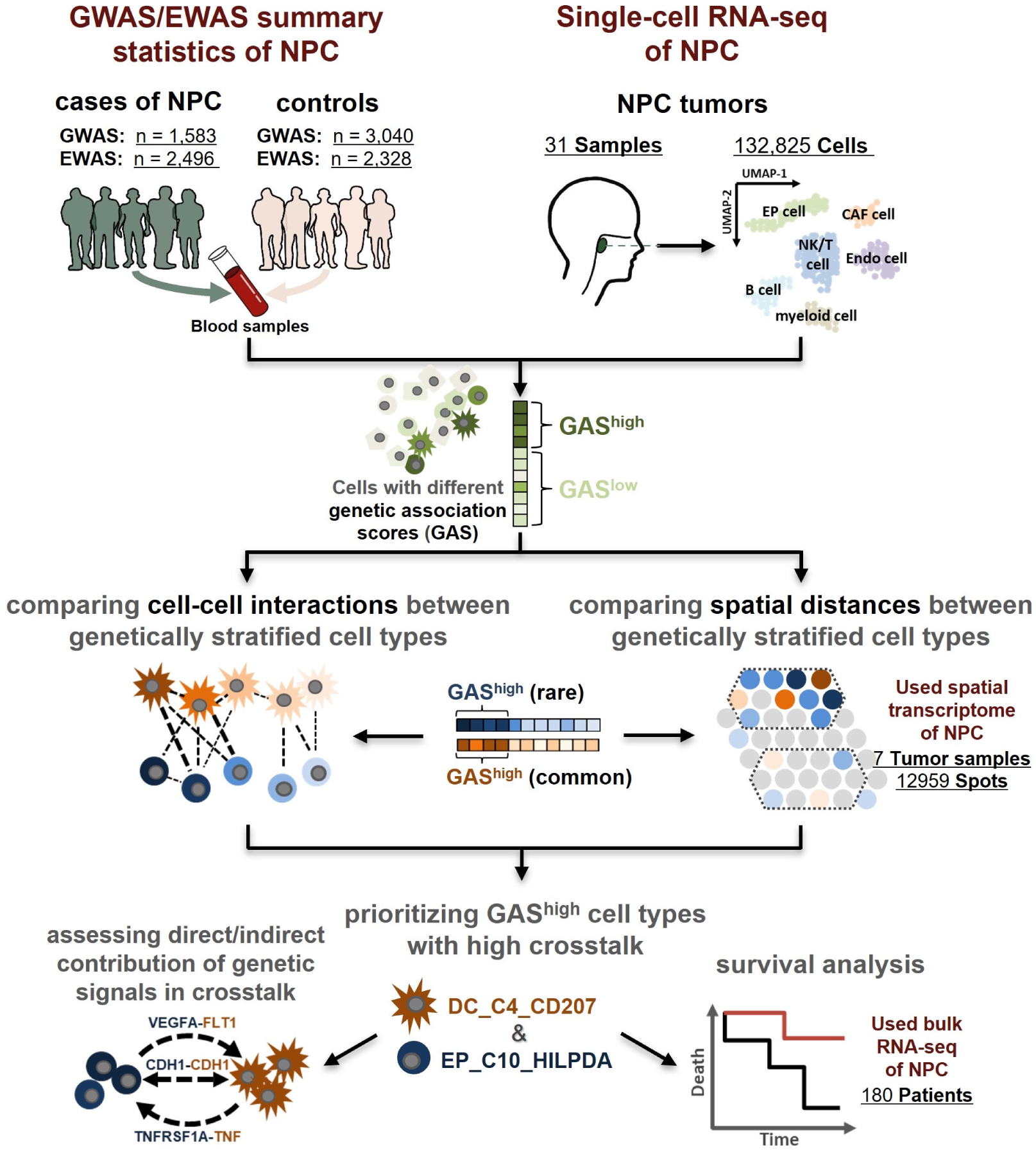
Schematic overview of study design. We integrated large-scale genetic association analyses of NPC, including common (genome-wide association study, GWAS; n_sample_=4,623) and rare variants (exome-wide association study, EWAS; n_sample_=4,824), with single-cell RNA sequencing (scRNA-seq) data of 31 NPC tumors as well as spatial transcriptomic data of 7 NPC tumors. Genetic Association Scores (GASs) were calculated for individual cells in scRNA-seq data, enabling the identification of cell subsets enriched for genetically associated genes. To prioritize functionally relevant cell populations, we compared genetically stratified cell types with respect to both cell-cell interactions and spatial proximity. Finally, functional assessments, leveraging scRNA-seq data from 31 NPC patients, and survival analyses, leveraging bulk RNA-seq data from 180 NPC patients, established the biological and clinical significance of prioritized cell populations and pathways.

## Results

### Genetic effects of common and rare variants are mediated through distinct cell types within NPC TME

To delineate the TME contexts mediating genetic risk for NPC, we first conducted gene-level association tests for both common (GWAS; n=4,623; MAF≥0.01) and rare variants (EWAS; n=4,824; MAF<0.01)^7,9,17^. EWAS was stratified into separate analyses of patients with NPC with family history (FHNPC) and sporadic NPC (SPNPC) to allow potential differences in the underlying mechanisms of rare variants (see Methods). We next integrated these association results with two independent scRNA-seq cohorts of NPC tumors (C1: 72,470 cells from 20 samples; C2: 60,355 cells from 11 samples; Fig. 1)^18–22^. Across both datasets, we identified 11 major cell types and 85 subtypes (**Fig. S1**). Using the scDRS framework^23^, we calculated cell-level genetic association scores (GASs) to quantify NPC-associated gene enrichment in individual cells and across cell populations.

This analysis revealed that 314 NPC-associated genes through common variants (P<0.005; **Supplementary Tables 1**) were significantly enriched in professional antigen presenting cells (APCs), particularly dendritic cells (DCs), monocyte-macrophage cells and B cells (**Fig. S2**). Among these, the DC_C4_CD207 subtype showed the strongest enrichment signal (**Fig. 2A-C**). Pathway-level analysis further confirmed that NPC-associated pathways through common variants (P_FDR_<0.05, **Supplementary Tables 2**) were preferentially active in DC_C4_CD207 (**Fig. 2D, Fig. S3**). By contrast, 269 NPC-associated genes through rare variants in FHNPC (P<0.005; **Supplementary Tables 1**) were significantly enriched in malignant epithelial cells (EPs), including EP_C10_HILPDA, EP_C7_MMP7, EP_C3_HLA and EP_C8_SLPI. In SPNPC, 186 NPC-associated genes through rare variants were enriched in inflammatory cancer-associated fibroblasts (iCAFs), particularly the iCAF_C1_MMP11 subtype (P_meta-Bonferroni_<0.05; **Fig. 2A-B; Fig. S2**). These enrichment patterns were highly consistent across both scRNA-seq cohorts (**Fig. 2A; Fig. S2, S4, S5**), collectively suggesting that common and rare genetic risk factors are linked with distinct cellular populations within the NPC TME.

**Figure 2.**
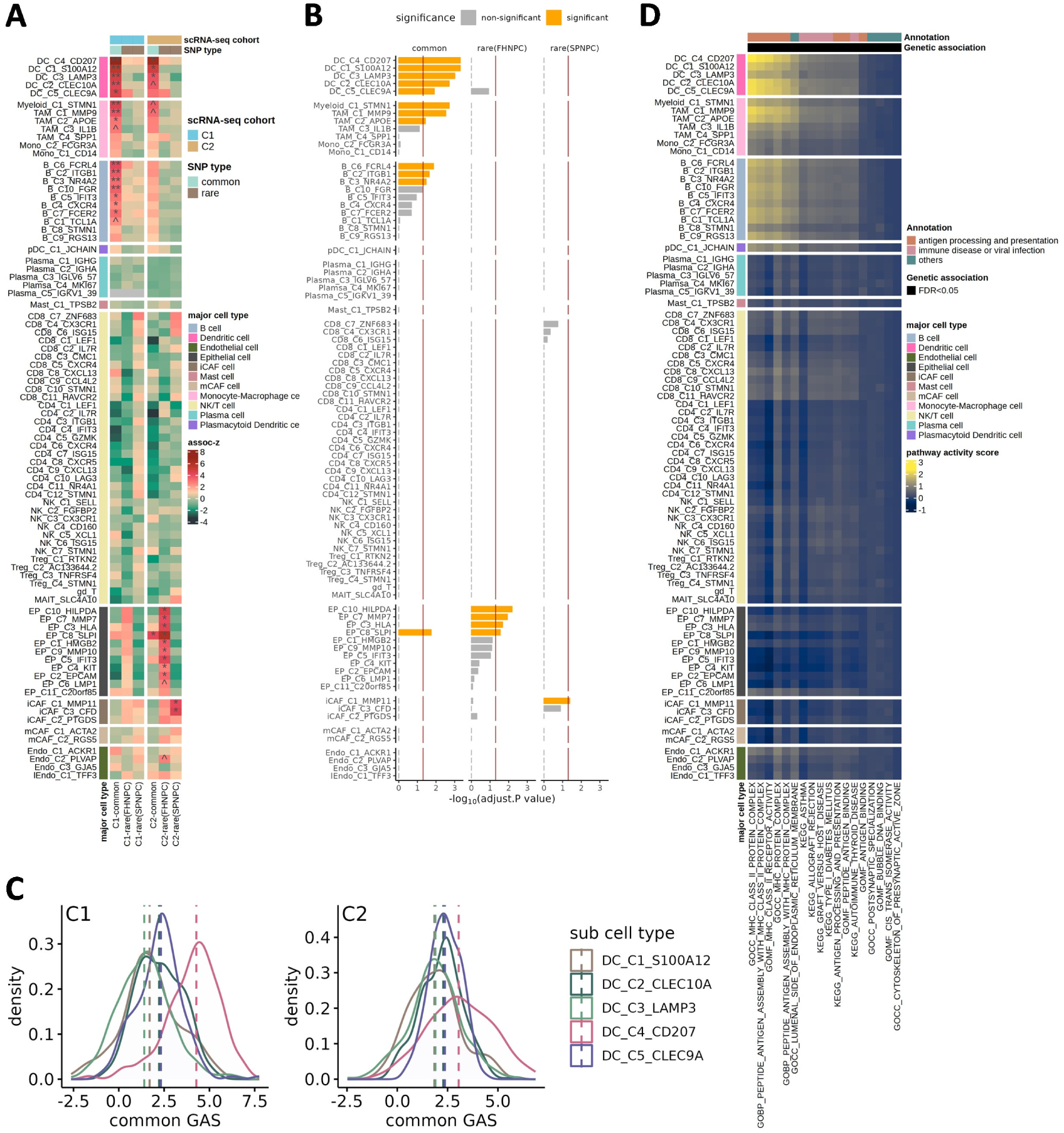
Identification of cell types mediating genetic effects in NPC TME. **A** Heatmap showing enrichment of NPC-associated genes’ expression across 85 cell types in two independent scRNA-seq cohorts (C1 and C2). Gene sets derived from common and rare variants were analyzed separately. Enrichment strength is represented by assoc-z values. P-values were adjusted using the FDR method. Statistical significance: ^, P<0.05; *, adjusted P<0.05; **, adjusted P<0.01. **B.** Meta-analysis summarizing enrichment results across both scRNA-seq cohorts for each cell subtype. P-values were adjusted using the Bonferroni method. Vertical red line indicates the significance threshold (adjusted P=0.05). **C.** Density plots illustrating the distribution of genetic association score (GAS) across dendritic cell (DC) subtypes in cohorts C1 and C2, highlighting consistently elevated scores in DC_C4_CD207. **D.** Heatmap of pathway activity scores for NPC-associated pathways (through common variants, adjusted P<0.05 with the FDR method) across 85 cell subtypes.

**Figure 3.**
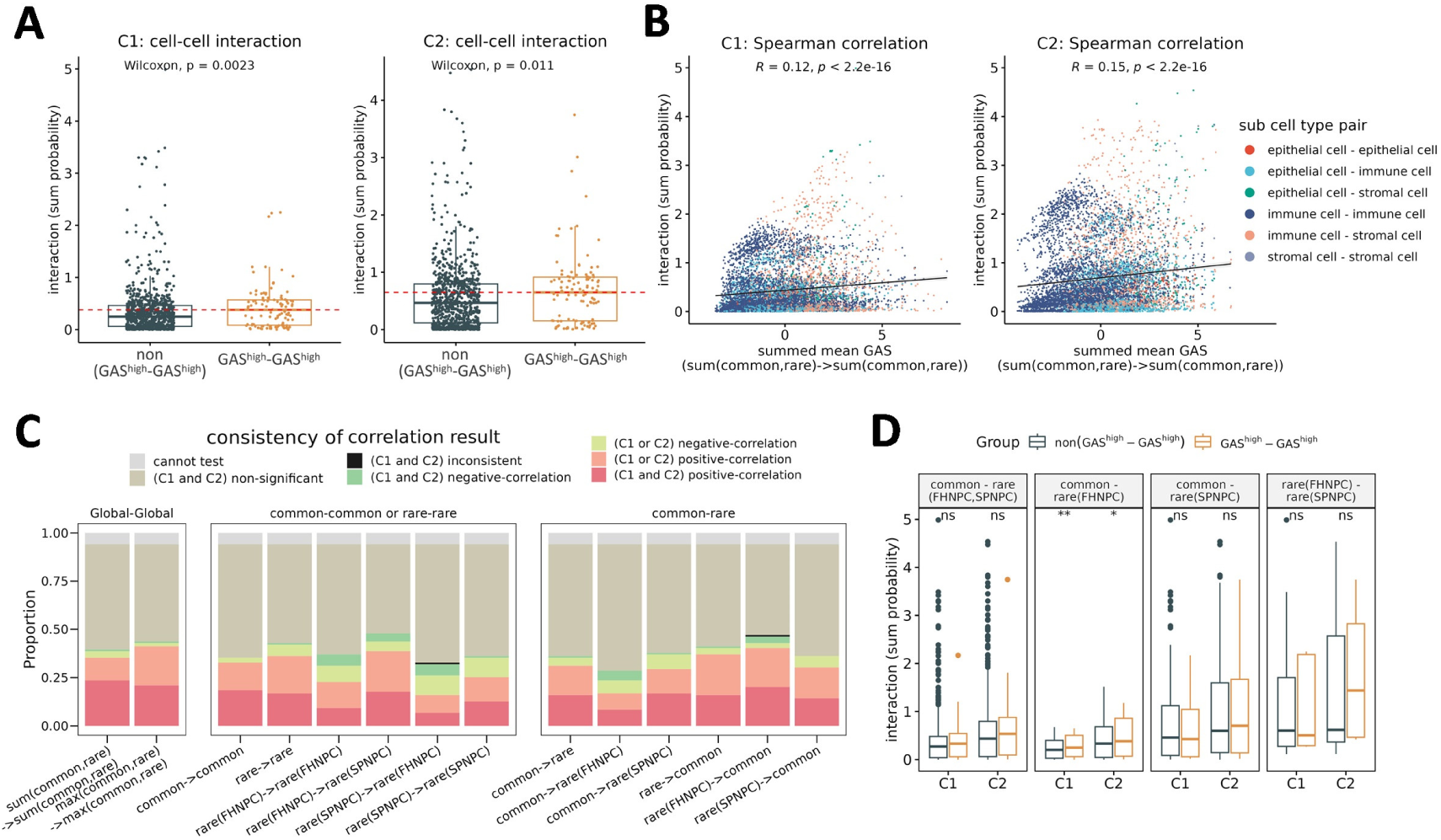
Enhanced cell-cell interactions among genetically associated cell types in NPC. **A** Boxplots showing cell-cell interaction strengths (summed interaction probabilities) between GAS^high^ - GAS^high^ versus other cell pairs in two independent scRNA-seq cohorts (C1 and C2). Statistical significance was assessed using Wilcoxon test. **B.** Scatter plots showing the correlation between summed mean GASs and the interaction strengths (summed interaction probabilities) across major epithelial, immune, and stromal cell-type pairs in the two cohorts. Cell types are color-coded as indicated. Correlations were assessed by Spearman test. **C.** Bar plots showing the between-cohort consistency of the correlation between the interaction strengths (summed interaction probabilities) and multiple GAS metrics, stratified by ligand-receptor directionality (e.g., common→rare). Colors indicate categories of correlation consistency between cohorts. **D.** Box plots comparing crosstalk-type-specific cell-cell interaction strengths across cell subtype pairing groups. GAS^high^ - GAS^high^ pairs for the “common-rare” type of crosstalk refer to cell pairs where one cell subtype is rated as GAS^high^ for common variant and the partner is rated as GAS^high^ for rare variants (FHNPC and/or SPNPC). Comparisons include GAS^high^ - GAS^high^ versus other GAS-GAS pairs not meeting the GAS^high^ - GAS^high^ criteria. Statistical significance was determined using Wilcoxon test: *, P < 0.05; **, P < 0.01; ns, not significant.

**Figure 4.**
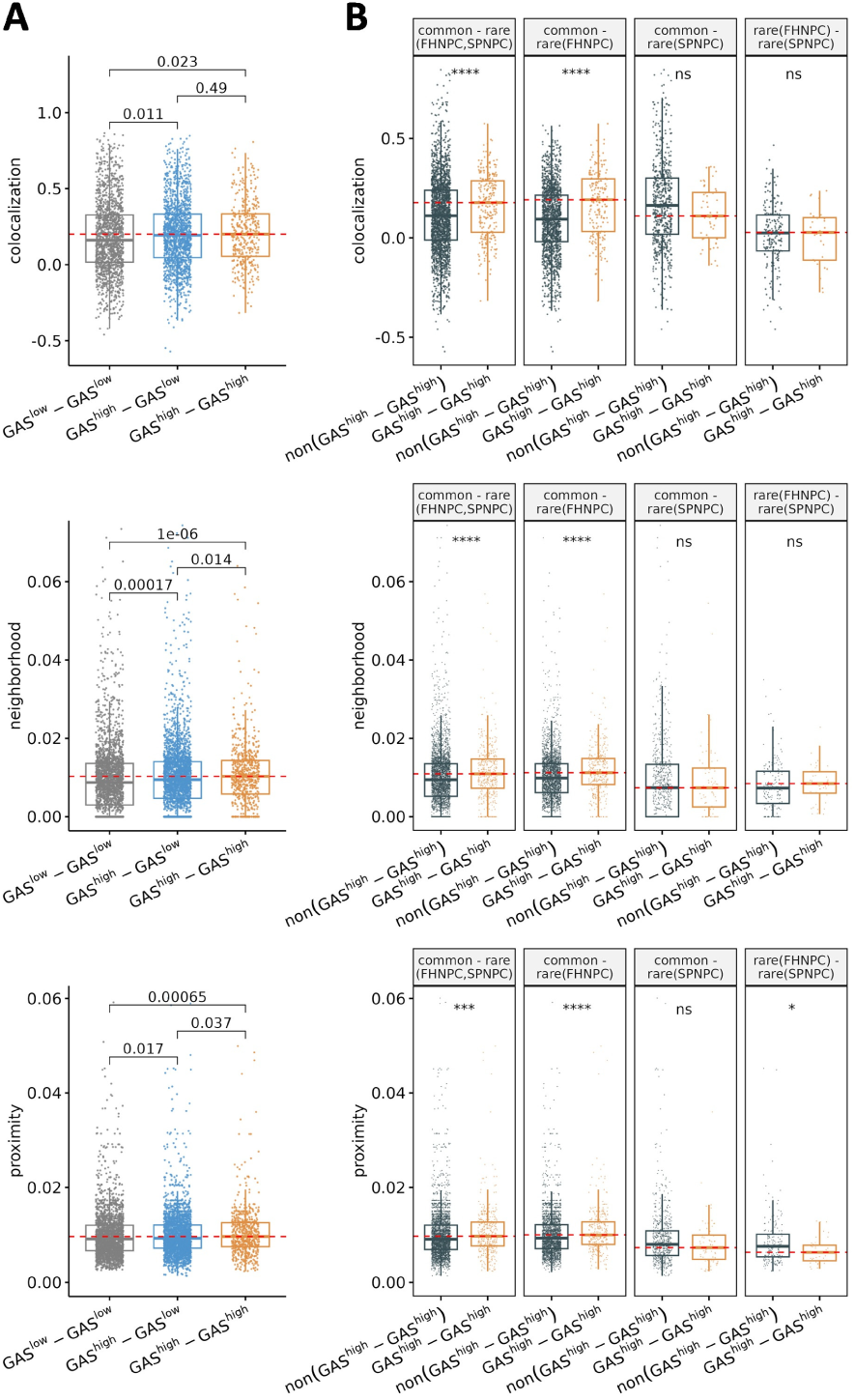
Spatial proximity of genetically enriched cell types within the NPC TME. **A** Box plots comparing overall spatial metrics (colocalization, neighborhood, and proximity scores) across three categories of cell subtype pairs: GAS^high^-GAS^high^, GAS^high^-GAS^low^ and GAS^low^-GAS^low^. Statistical testing was performed using the Wilcoxon test. **B.** Box plots showing crosstalk-type-specific comparisons of spatial metrics. GAS^high^ - GAS^high^ pairs for the “common-rare” type of crosstalk refer to cell pairs where one subtype is rated as GAS^high^ for common variant and the partner is rated as GAS^high^ for rare variants (FHNPC and/or SPNPC). Comparisons include GAS^high^ - GAS^high^ versus other GAS-GAS pairs not meeting the GAS^high^ - GAS^high^ criteria. Statistical significance was determined using Wilcoxon test: *, P < 0.05; **, P < 0.01; ***, P < 0.001; ****, P < 0.0001; ns, not significant.

**Figure 5.**
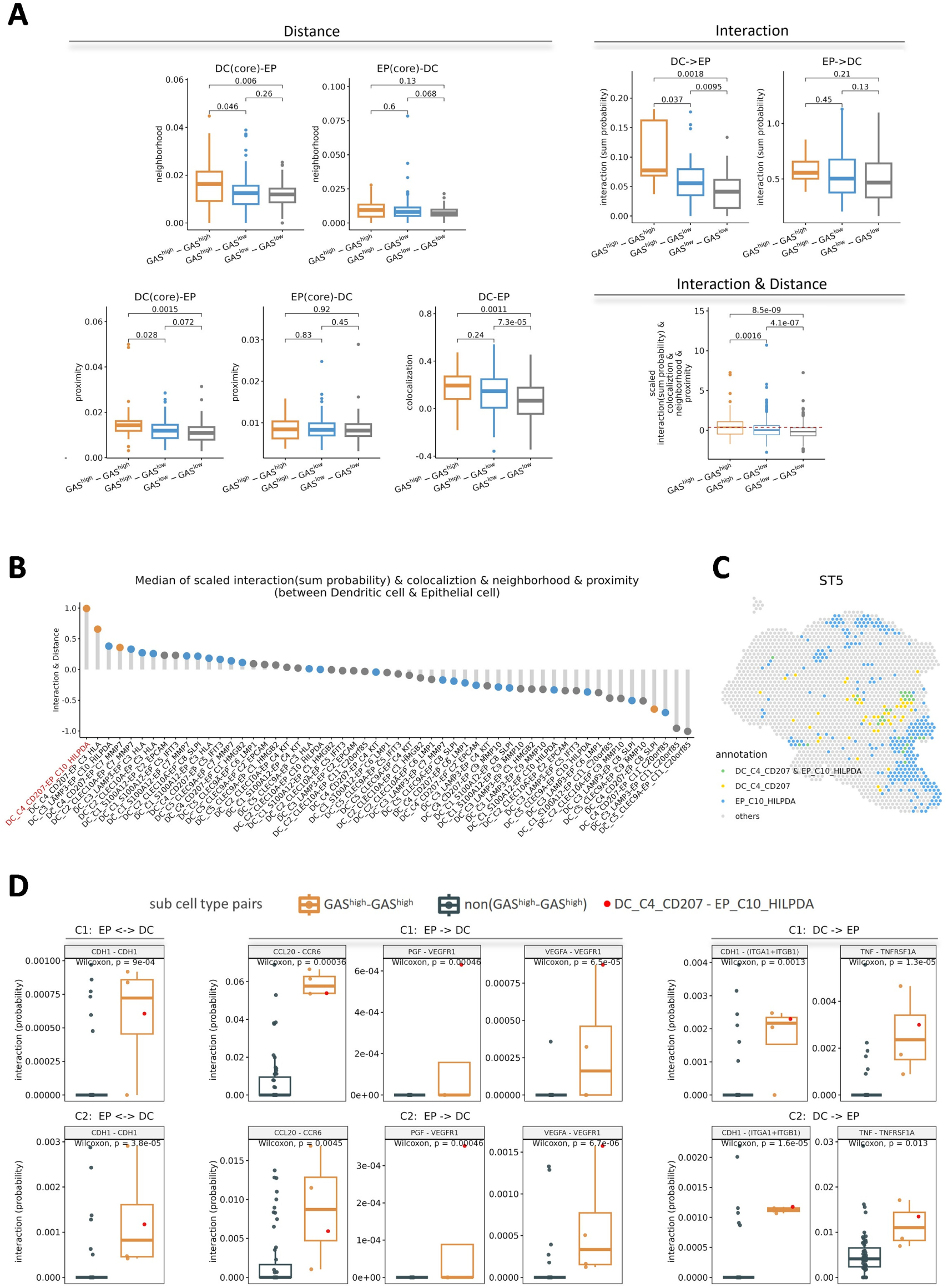
Strong intercellular interaction and spatial proximity between dendritic DC_C4_CD207 and epithelial EP_C10_HILPDA cells. **A** Box plots comparing interaction probability (labeled as “Interaction”), spatial proximity scores (labeled as “Distance”), and their combined metric (labeled as “Interaction & Distance”) across dendritic-epithelial cell subtype pairs grouped by GAS categories (GAS^high^-GAS^high^, GAS^high^-GAS^low^ and GAS^low^-GAS^low^). Wilcoxon tests were used to assess significance. **B.** Ranked dot plot of integrated interaction and spatial measurement (“Interaction & Distance”, involving sum probability of cell-cell interaction, colocalization, neighborhood, proximity) across all dendritic-epithelial subtype pairs, highlighting DC_C4_CD207-EP_C10_HILPDA as the top-ranked pair (red). **C.** Representative visualization of NPC ST data (sample ST5), showing ST spots containing DC_C4_CD207 only (yellow), EP_C10_HILPDA only (blue), or both subtypes (green). **D.** Box plots of six ligand(L)-receptor(R) pairs where interaction probability in “GAS^high^ - GAS^high^” cell subtype pairs of the crosstalk between EP and DC was significantly higher than in “non(GAS^high^ - GAS^high^)” pairs. Each dot represents an interaction probability in a specific subtype pair; DC_C4_CD207-EP_C10_HILPDA subtype pair is highlighted in red. EP → DC and DC → EP denote interaction directionality from ligand-expressing to receptor-expressing cells; ↔ indicates bidirectional interactions. DC(core) and EP(core) refer to spatial proximity metrics centered on dendritic or epithelial cell types, respectively. Significant difference was determined using Wilcoxon test.

### Cell types enriched for genetic associations exhibit stronger interactions within the NPC TME

Given the critical role of TME crosstalk in cancer development^24,25^, we next explored whether genetic variation influence cell-cell interactions. Across both scRNA-seq cohorts, interaction strengths were significantly higher between cell subtypes with high GAS (GAS^high^-GAS^high^) compared to those GAS^high^-GAS^low^ or GAS^low^-GAS^low^ pairs (**Fig. 3A, Fig. S6A**). Moreover, interaction strength positively correlated with the summed genetic association burden (summed mean GAS of the interacting cell type pairs) across epithelial, immune, and stromal cells (Spearmen R =0.12 for C1; R=0.15 for C2; P<2.2×10^-16^; **Fig. 3B**). These correlations were robust across different GAS metrics considering allele frequency and crosstalk-types (**Fig. 3C**), and consistent in sample-level analyses (**Fig. S7**). Together, these findings support a generalizable link between genetic associations and intercellular communication within the TME.

We then focused on a specific type of crosstalk - those between a cell subtype with high GAS for common variant (GAS^high(common)^) and the partner cell subtype with high GAS for rare variants (GAS ^high(rare)^). The GAS^high(common)^-GAS^high(rare)^ pairs exhibited significantly stronger interactions than GAS^low(common)^-GAS^low(rare)^ or mixed pairs, particularly when rare variants were derived from FHNPC (**Fig. 3D, Fig. S6B**). This enhancement of interactions was not observed for rare variants identified in SPNPC (**Fig. 3D, Fig. S6B**). Collectively, these findings suggest that genetically enriched cell populations not only mediate individual-variant-level risk but also engage in preferential crosstalk, pointing to germline variants as modulators of intercellular communication network in NPC development.

### Cell types enriched for genetic associations exhibit closer spatial proximity within the NPC TME

Considering GAS^high^cell types have stronger intercellular interactions, we hypothesized that these populations may also reside in closer physical proximity within the TME. To test this, we analyzed spatial transcriptome (ST) data from seven NPC tumors (**Fig. 1**). Each sample contained an average of 1,852 spots, which was deconvoluted into 3-7 cell subtypes using RCTD with a permutation-based approach^26^ (see Methods; **Fig. S8A-B**). Spatial proximity between cell subtypes was assessed using colocalization, neighborhood, and proximity scores (see Methods). Across all metrics, GAS^high^-GAS^high^ pairs consistently demonstrated significantly higher spatial proximity compared to GAS^high^-GAS^low^ and GAS^low^-GAS^low^ pairs (**Fig. 4A**). Notably, GAS^high(common)^-GAS^high(rare)^ pairs exhibited elevated spatial proximity scores, especially when rare variants were derived from FHNPC (**Fig. 4B**). These findings collectively demonstrate that genetically enriched cell populations are preferentially positioned in close spatial neighborhoods, thereby facilitating intercellular communication that may underlie NPC tumorigenesis.

### Significant spatial crosstalk between dendritic cell subtype DC_C4_CD207 and epithelial cell subtype EP_C10_HILPDA

Since dendritic and epithelial cells showed the strongest enrichment for common and rare variant associations, respectively (**Fig. 2**), we next investigated their intercellular crosstalk. We observed that unidirectional dendritic-to-epithelial interactions (“DC→EP”) were significantly elevated in GAS^high^-GAS^high^ subtype pairs compared to GAS^high^-GAS^low^ and GAS^low^-GAS^low^ pairs (**Fig. 5A, Fig. S9**). Bidirectional ligand-receptor (L-R) interactions between dendritic and epithelial cells were also significantly stronger in GAS^high^-GAS^high^ pairs, suggesting enhanced mutual communications among genetically enriched subtypes (**Fig. S10, Fig. S9**). Spatial analyses further demonstrated that dendritic cells and adjacent epithelial cells (labeled as “DC(core)-EP”), specifically those involving GAS^high^ subtypes, exhibited significantly higher colocalization, neighborhood, and proximity scores (**Fig. 5A, Fig. S9**), supporting tighter spatial organization of these populations within the TME.

When jointly assessing both interaction probability and spatial proximity, the DC_C4_CD207-EP_C10_HILPDA pair ranked as the strongest dendritic-epithelial crosstalk partner across all subtype combinations (see Methods; **Fig. 5B, Fig. S11**). Meta-analysis of spatial transcriptomics data across samples confirmed frequent colocalization between these two subtypes (**Fig. 5C, Fig. S12**), which were also among the most genetically enriched populations (**Fig. 2A-B**).

To elucidate the molecular drivers of this interaction, we identified 20 ligand-receptor (LR)-pairs with significant interaction probabilities (P<0.05), of which 11 were preferentially enriched in the DC_C4_CD207-EP_C10_HILPDA pair (P_FDR_<0.05, **Fig. S13**). Notably, six LR-pairs (CDH1-CDH1, CCL20-CCR6, PGF-VEGFR1(FLT1), VEGFA-VEGFR1(FLT1), CDH1-(ITGA1+ITGB1), TNF-TNFRSF1A) also showed overall enriched interaction across GAS^high^-GAS^high^ subtype pairs relative to GAS^low^-GAS^low^ cell type pairs (**Fig. 5D**). These LR axes may thus represent molecular anchor points mediating genetically driven spatial crosstalk between key immune and epithelial compartments in NPC pathogenesis.

### Direct and indirect contribution of genetic susceptibility to the DC_C4_CD207 and EP_C10_HILPDA crosstalk

To investigate how genetic susceptibility contributes to communications between DC_C4_CD207 and EP_C10_HILPDA, we first questioned whether NPC-associated genes directly encode ligands or receptors mediating their interaction. Using Fisher’s exact test, we observed a significant overlap between common-variant-mediated NPC-associated risk genes and ligand genes expressed in DC_C4_CD207, including multiple HLA family members (**Fig. 6A-B, Fig. S14**), which play crucial role in various cancers including NPC^27–29^. Sensitivity analysis confirmed that HLA genes broadly contributed to such genetic enrichment across many cell types (**Supplementary Tables 3, Fig. S15**). While after excluding HLA genes in the NPC-associated gene set, only LR encoding genes expressed in DC_C4_CD207 and B_C8_STMN1 subtypes remained significant across both scRNA-seq cohorts (**Supplementary Tables 3**), underscoring the specific involvement of DC_C4_CD207 in NPC-associated LR crosstalk. As supporting evidence, non-HLA NPC susceptibility genes such as TNF (P_common_ = 2.173×10^-5^) and F11R (P_common_ = 0.024) also encode ligands directly implicated in DC_C4_CD207-EP_C10_HILPDA interactions (**Fig. 6B**).

**Figure 6.**
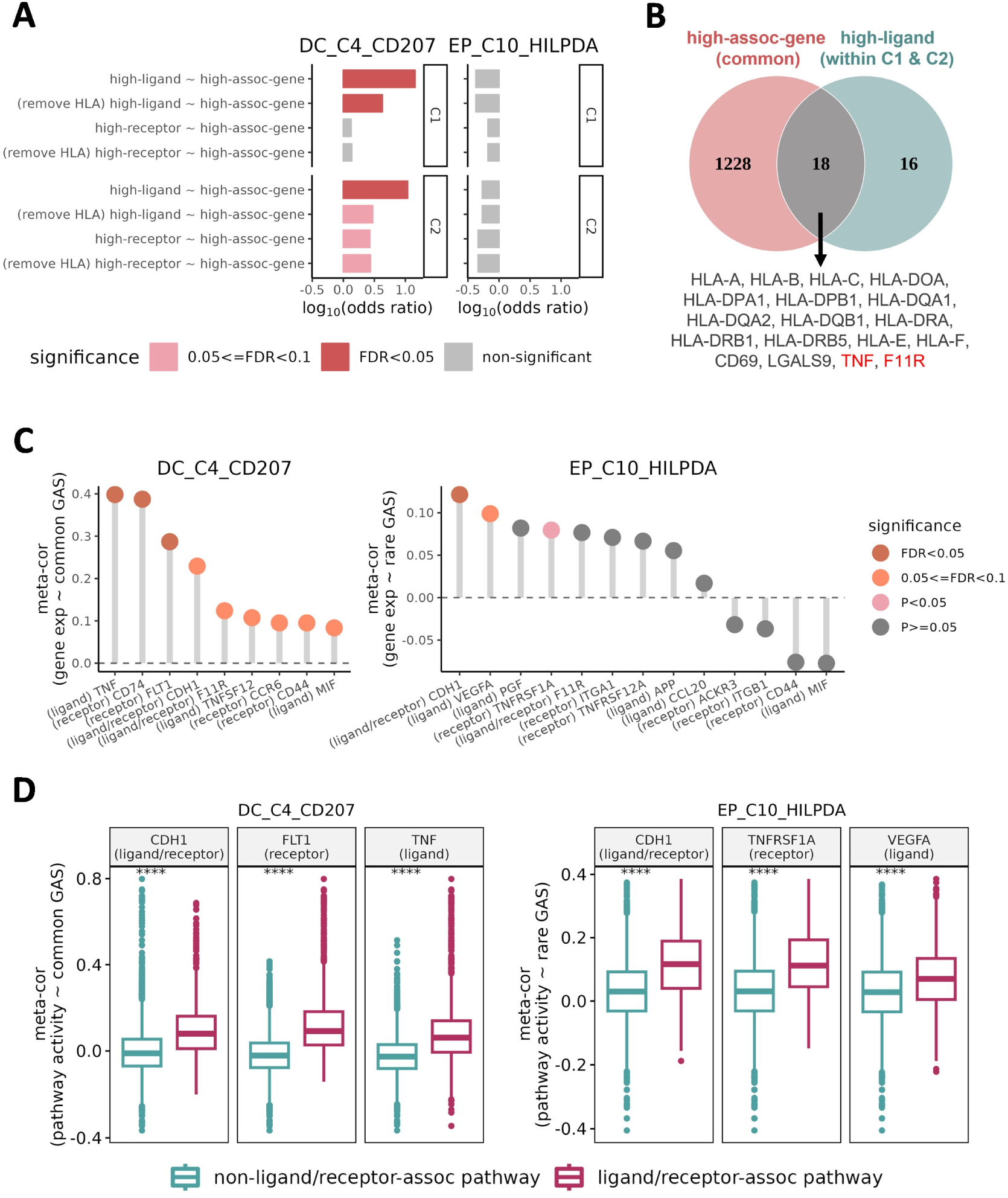
Direct and indirect involvement of NPC-associated susceptibility genes in the DC_C4_CD207 and EP_C10_HILPDA crosstalk. **A** Bar plot showing odds ratios for enrichment of NPC-associated susceptibility genes (P<0.05), detected using common (for DC_C4_CD207) or rare (for EP_C10_HILPDA) variants, among ligands (L) or receptors (R) encoding genes in DC_C4_CD207 and EP_C10_HILPDA. P values from Fisher’s exact test were corrected by FDR, significant results were shaded in red. **B.** Venn diagram showing overlap between NPC-associated susceptibility genes (through common variants) and ligand genes involved in significant interacting LR pairs (P<0.05) in DC_C4_CD207 (in both C1&C2 cohorts). Genes highlighted in red directly contribute to crosstalk with EP_C10_HILPDA. **C.** Ranked dot plots showing meta-analyzed Spearman correlations between GAS and expression of key LR genes in DC_C4_CD207 (left) and EP_C10_HILPDA (right). Colors denote significance after FDR correction. **D.** Box plots comparing meta-analyzed Spearman correlations between GAS and pathway activation scores for LR-associated versus non-LR-associated pathways in DC_C4_CD207 and EP_C10_HILPDA. Examples shown for CDH1(a ligand/receptor, left panel), FLT1(a receptor, middle panel), TNF (a ligand, right panel), TNFRSF1A (a receptor, middle panel), and VEGFA (a ligand, right panel). Wilcoxon tests: ****, P < 0.0001. C1: cohort 1, C2: cohort 2. meta-cor: meta-analyzed Spearman correlation coefficient; common GAS: GAS derived from common variants; rare (FHNPC) GAS: GAS derived from rare variants in FHNPC patients.

We next hypothesized that genetic susceptibility may also influence intercellular communication indirectly by modulating LR-associated pathways. Among the 11 key LR-pairs with significantly elevated interaction probability in the DC_C4_CD207 - EP_C10_HILPDA axis (**Fig. S13**), expression of TNF-TNFRSF1A, VEGFA-VEGFR1(FLT1), and CDH1-CDH1 was significantly correlated with GAS in DC_C4_CD207 (common variants) or in EP_C10_HILPDA (rare variants from FHNPC; **Fig. 6C**). Notably, only TNF itself was included in the GAS calculation, suggesting that other LR genes may be indirectly regulated through downstream pathways. Consistent with this, analysis of 12,885 curated molecular pathways^30^ revealed that LR-associated pathways (e.g., those linked to TNF, FLT1 and CDH1 in DC_C4_CD207, and TNFRSF1A, VEGFA and CDH1 in EP_C10_HILPDA) showed significantly stronger correlations with GAS compared to non-LR-associated pathways in both cell types (P<2.2×10^-16^; **Fig. 6D**). These pathways also overlapped significantly with GAS-associated pathways and NPC genetically associated pathways identified through gene set–level association tests (see Methods; **Fig. S16**). Collectively, these findings demonstrate NPC genetic susceptibility influences the DC_C4_CD207-EP_C10_HILPDA axis through both direct mechanisms, where susceptibility genes encode key LR molecules, and indirect mechanisms, where susceptibility-associated pathways regulate LR-mediated signaling, particularly involving TNF–TNFRSF1A, VEGFA–VEGFR1, and CDH1–CDH1interactions.

### Biological and clinical relevance of DC_C4_CD207 and EP_C10_HILPDA in NPC

To characterize the biological functions of DC_C4_CD207 and EP_C10_HILPDA, we identified featured pathways distinguishing each subtype from other dendritic and epithelial subpopulations (see Methods). For DC_C4_CD207, the featured pathways mainly involved antigen presentation and immune activation functions (**Fig. S17, S18**). Marker genes such as CD1A and HLA-DQA2 supported antigen presentation roles^31,32^, while CLEC4A and IL18 implicated immune signaling and regulation (**Fig. S19)**^33,34^.

For EP_C10_HILPDA, featured pathways highlighted hypoxia response and metabolic reprogramming, including glycolysis (**Fig. S17, S18**). Consistent with this, marker genes such as ENO1 and LDHA indicated glycolytic activity^35,36^, while HILPDA and BNIP3 reflected hypoxia adaptation^37,38^ (**Fig. S19**). Notably, the upregulated pathways in both subtypes significantly overlapped with their LR-associated pathways, suggesting that genetically-regulated intercellular communications form at least partially the molecular programs defining these cells (**Fig. S16**).

We next evaluated their clinical relevance using infiltration estimates from 180 bulk NPC tumor samples with ssGSEA^39^. Cox regression analyses revealed that higher infiltration of DC_C4_CD207 was significantly associated with improved disease-free survival (DFS; HR= 0.646, P= 0.037, R^2^=0.171), while higher infiltration of EP_C10_HILPDA predicted worse DFS (HR=2.448, P=0.022, R^2^=0.207) (**Supplementary Tables 4**). Kaplan-Meier (K-M) curve survival analyses confirmed that patients in the lowest quartile (bottom 25%) for DC_C4_CD207 infiltration or the highest quartile (top 25%) for EP_C10_HILPDA infiltration had poorer DFS (P_DC_C4_CD207_ =0.001, P_EP_C10_HILPDA_ =0.061, log-rank; **Fig. 7A**), with consistent results under alternative thresholds (**Fig. S20**). A multivariable Cox regression model incorporating both cell types provided greater predictive power (R^2^=0.309) compared to single-cell-type models or other epithelial/dendritic subtypes (**Fig. 7B, Supplementary Tables 4, Fig. S21A**). Patients with high EP_C10_HILPDA but low DC_C4_CD207 infiltration had the worst DFS (**Fig. 7C**). Conversely, a higher DC_C4_CD207 to EP_C10_HILPDA infiltration ratio was associated with favorable DFS (**Fig. 7B, Supplementary Tables 4, Fig. S21B**). Collectively, these findings suggest that DC_C4_CD207 and EP_C10_HILPDA represent biologically distinct yet clinically relevant cell populations in NPC progression and exhibit promising potential to stratify NPC patients with distinct prognosis.

**Figure 7.**
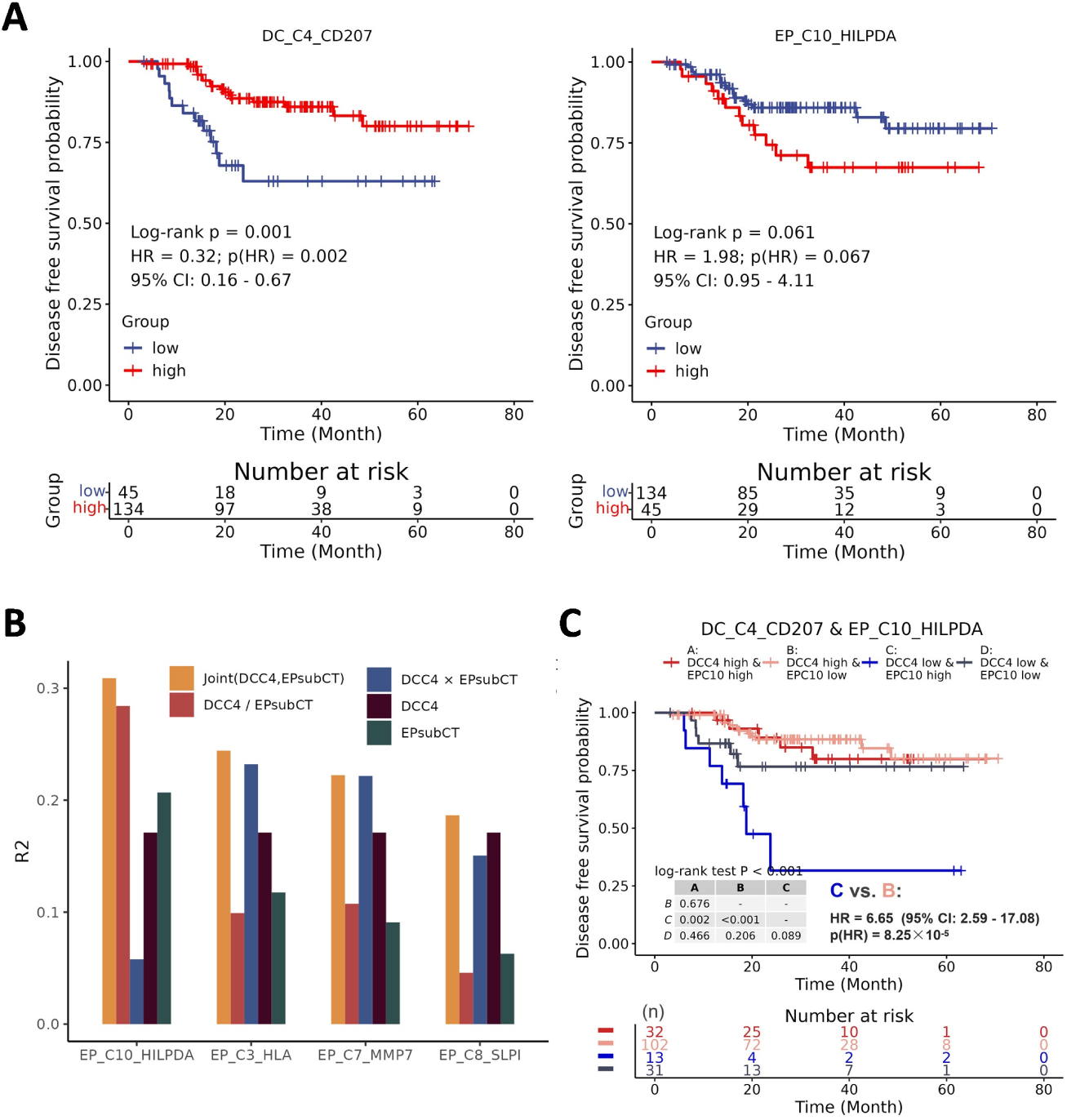
Clinical relevance of DC_C4_CD207 and EP_C10_HILPDA infiltration in NPC. **A** Kaplan-Meier (KM) survival curves of disease-free survival (DFS) stratified by infiltration levels of DC_C4_CD207 (left) or EP_C10_HILPDA (right). Survival differences were compared using log-rank tests: patients in the bottom 25% vs top 75% for DC_C4_CD207 infiltration (left, low infiltration corresponds to worse DFS), and patients in the top 25% vs bottom 75% for EP_C10_HILPDA infiltration (right, higher infiltration corresponds to worse DFS). **B.** Bar plot presenting performance (R^2^) of target term in Cox regression model across different modeling strategies: DC_C4_CD207 (DCC4, univariate); epithelial subtype (EPsubCT, univariate); additive model (DCC4 + EPsubCT); interaction term (DCC4 × EPsubCT) in interaction model; and infiltration ratio (DCC4 / EPsubCT). **C.** KM curves stratifying patients into four groups based on high/low infiltration (same cut-points as defined in **A**) of DC_C4_CD207 and EP_C10_HILPDA. Patients with low DC_C4_CD207 and high EP_C10_HILPDA infiltration exhibited the poorest DFS. For all KM analyses, the 25th or 75th percentiles were used as cut-points for infiltration levels of DC_C4_CD207 or EP_C10_HILPDA, respectively. Hazard ratios (HR) and 95% CIs were estimated by Cox regression.

## Discussion

By integrating genetic association data with single-cell, spatial, and bulk transcriptomics, we reveal that germline variants in NPC shape tumor biology not soly, but by synergistically structuring networks of intercellular communication within the TME. Common variant associations mapped predominantly to APCs, while rare variant associations localized to malignant epithelial and stromal subsets (iCAFs). These genetically enriched cell populations manifested enhanced intercellular interactions and closer spatial proximity, with the strongest crosstalk observed between Langerhans dendritic cells (DC_C4_CD207; enriched for common variants) and and HILPDA⁺ epithelial cells (EP_C10_HILPDA; enriched for rare variants). Joint consideration of these subtypes markedly improved prognosis prediction, highlighting the clinical significance of genetically defined TME networks.

These findings align with the model in which common low-penetrance variants establish baseline liability, while rare high-penetrance variants trigger disease onset^40^. In NPC, common variants appear to predispose immune dysfunction, while rare variants directly affect tumor-originating epithelial cells. Functionally, these processes converge, as impaired antigen presentation facilitates tumor progression^41^. Familial NPC, known for earlier onset and more aggressive disease^42,43^, was enriched for rare-variant-defined genes in malignant epithelial cells, implicating rare epithelial variants as drivers of familial clustering. This suggests that differences in cellular context of genetic risk may underlie distinct clinical subtypes of cancer.

Our results propose a new paradigm: assessing germline risk through the lens of intercellular crosstalk. Cell type pairs enriched for common and rare variant risk consistently exhibited stronger interactions and spatial proximity, providing evidence that germline polygenicity converge at the intercellular level. This complements previous studies focusing on epistasis or intracellular protein interactions^44–46^, and reframes polygenic risk as a network-level phenomenon.

Mechanistically, the DC_C4_CD207 and EP_C10_HILPDA emerged as the strongest genetically defined interaction. Langerhans cells are specialized APCs within epithelial tissues^47,48^, with known functions in initiating immune responses against cancer antigens^49,50^. However, their role in NPC remains unclear. We identified VEGFA-FLT1 signaling from EP_C10_HILPDA to DC_C4_CD207 as a key pathway, alongside TNF-TNFRSF1A and CDH1-CDH1 interactions, ligand-receptor pairs genetically implicated either directly or through risk-associated pathways. Previously, studies have reported roles of VEGFA-FLT1 in immune suppression and angiogenesis function^51,52^, TNF-TNFRSF1A in inflammatory signaling function ^53,54^, and CDH1-CDH1 in intercellular adhesion function^55^. Our data, together with existing evidence, suggest that germline susceptibility both encodes and regulates intercellular signaling axes linking epithelial and immune compartments in TME.

Our clinical data supported that infiltration of DC_C4_CD207 or EP_C10_HILPDA alone stratifies patient survival, aligning with established associations of Langerhans cells with favorable prognosis in NPC and high tumor HILPDA with poor outcomes in cancers^56–59^. Moreover, joint modeling of both subtypes yielded greater prognostic power, indicating that genetically defined crosstalk, rather than single populations, may provide more accurate biomarkers. Such interactions may also represent therapeutic entry points, particularly in strategies aimed at reprogramming TME communication.

Our study has limitations. SNP-gene assignment relied on proximity, and functionally informed mapping (e.g., Hi-C, multi-OMIC QTL) may reveal additional targets. Associations do not establish causality, necessitating experimental validation of candidate interactions. Finally, while NPC serves as a model, testing the generalizability of these findings across other polygenic cancers and complex diseases will be important.

### Conclusions

We establish a paradigm in which germline variants influence cancer through genetically defined intercellular crosstalk. In NPC, common-variant–associated APCs and rare-variant–associated epithelial cells engage in stronger, spatially proximate interactions, with the DC_C4_CD207–EP_C10_HILPDA axis representing a central convergence point. These interactions are mediated by ligand-receptor pairs genetically linked to NPC risk and have prognostic value when considered jointly. More broadly, our results highlight intercellular communication as a critical mechanism by which germline polygenicity shapes tumor biology, with implications for biomarker development and therapeutic targeting.

## Materials and Methods

### Datasets

#### Genome-wide association study (GWAS) and exome-wide association study (EWAS) datasets

GWAS and EWAS datasets were obtained from published studies. Specifically, the GWAS dataset contained SNP-level association results of 5,503,891 variants with minor allele frequency ≥ 0.01 by analyzing 1,583 cases and 3,040 controls of southern Chinese descent after quality control filtering. Details of data generation and processing have been previously described^7,17^. The EWAS dataset contained whole-exome sequencing data of blood samples from 2,694 NPC patients and 2,328 controls (3,039 NPC patients and 2,692 controls before quality control) of southern Chinese descent recruited from Guangdong, China (N_npc_=2,125, N_control_=1,068) and Singapore (N_npc_=569, N_control_=1,260) as previously described^9^. Among all NPC cases in EWAS dataset, family history information (presence or absence) was available for 2,496 cases. Based on this, we obtained EWAS results from separate gene-based association tests performed for NPC patients with a family history (FHNPC, N_npc_=408) or without (sporadic NPC cases, SPNPC, N_npc_=2,088) to allow varied rare variants clustered in familial and sporadic cases.

#### Single-cell RNA-seq datasets

The scRNA-seq datasets were collected and processed in our previous study^4^. Details about scRNA-seq data generation and preprocessing, including sequencing, alignment, counting and normalization have been described previously^4^. The initial collection included data from 56 samples. EBV infection in epithelial cells have been considered to be an crucial event in the TME of NPC^18,60,61^, but the capture rate of epithelial cells infected with EBV varied across samples^4,9,20^. Therefore, we only retained 31 samples with EBV^+^ epithelial cells successfully captured in downstream analysis. Among those samples, twenty were sequenced using 3’-end RNA sequencing (labeled as “C1” cohort, N_cell_=72,470)^20–22^ and eleven was sequenced using 5’-end RNA sequencing (labeled as “C2” cohort, N_cell_=60,355)^18,19^. Single-cell-level expression matrix (N_sample_=31, N_cell_=132,825) and cell annotations were obtained in our previous study^4^. Annotation of sub cell types have been described previously^4^and were briefly summarized here: 1) removing the potential batch effect with Harmony algorithm^62^, 2) dimensionality reduction and clustering with R function “RunUMAP” and “FindClusters” in Seurat^63^, 3) using well-known markers to annotate cell clusters based on average gene expression and well-known cell type specific markers, 4) removing cell clusters overexpressing markers of two or more cell types, and 5) for each cell cluster extracting data of all annotated cells and repeating abovementioned step 1) - 4) to annotate sub cell clusters (sub cell type).To facilitate downstream analysis, the annotated sub cell types were projected to major cell types based on average gene expression profiles. Specifically, we utilized the R package Seurat V.4.3.0^63^ to calculate the average expression of sub cell types and selected the top 50 marker genes with specificity in expression for each sub cell type. Ribosomal protein (RPL/RPS) genes, mitochondrial genes, and immunoglobulin genes were excluded from the marker genes. The average expression profiles of marker genes for each sub cell type were used to calculate the Pearson correlation between pairwise cell types. The distance between pairwise sub cell types, calculated as 1 minus the Pearson correlation coefficient, was used in hierarchical clustering based on the “complete” method. Annotated major cell types included epithelial cells, DCs, endothelial cells, iCAFs, mCAFs, B cells, Plasma cells, pDC, NK/T cells, Monocyte-Macrophage cells, Mast cells (**Fig. S1**).

#### Bulk RNA-seq datasets

We analyzed bulk RNA-seq data of 180 NPC tumor samples collected and processed from previous study^4^. Details about its generation and processing have been described previously^4^. The datasets included samples from two cohorts. The bei-cohort consisted of RNA-seq data from tumor biopsies in the nasopharynx before any treatment from 92 NPC patients with prognostic data available^18^. The zhang-cohort consisted of RNA-seq data from tumor tissues of 88 NPC patients with prognostic data available^64^. Transcripts per million (TPM) were used in downstream analysis.

#### Spatial transcriptome datasets

Data generation: we analyzed spatial transcriptomic data of seven NPC tumor tissue samples. Data collection and pre-processing have been described previously^4^. In brief, The Visium Spatial Gene Expression for FFPE workflow (10X Genomics) were used for spatial transcriptome detection, generation and preprocessing of seven NPC FFPE tumor samples. We imported these Visium data of seven NPC patients via Seurat V.4.3.0^63^ for downstream analysis.

Cell type deconvolution of spatial transcriptome: the Visium spatial transcriptome data contains expression data at the spot level, with each spot covering an area of approximately 55 micrometers (μm) and potentially containing 1 to 10 cells. We utilized the tool RCTD (Robust cell type decomposition)^26^ in R package spacexr (version 2.2.0) to map all sub cell types captured and annotated from scRNA-seq data onto the spatial transcriptome. The specific processing pipeline of RCTD is as follows: The entire scRNA-seq expression matrix and sub cell type annotations are inputted as references. The “full” model is then used to perform deconvolution of the spatial transcriptome data, the process only applied to sub cell types with a cell count greater than or equal to 10 in the scRNA-seq data. Through deconvolution of 12,959 spots from seven samples after filtering with default RCTD parameters, we estimated the proportions of each sub cell type within each spot.

Additionally, to determine whether a specific cell type exists in a given spot of the spatial transcriptome sample, we computed a positive threshold based on the cell-type-specific proportions (positivity indicates the presence of a specific cell type in the spot). If the proportion of a cell type within a spot exceeded this positive threshold, we considered that cell type to be present in that spot. The threshold was cell-type specific and calculated through the following steps. We first generated multiple (n > 130) permuted spatial transcriptome matrixes with gene names randomly shuffled using the R function “sample”, and then repeated the deconvolution process using RCTD multiple times with these shuffled matrixes. Thus, for each sub cell type, we obtained multiple (n > 130) “pseudo” estimated proportions in each spot. We observed that when the number of random deconvolution runs reached 130, the “pseudo” estimated proportions for each sub cell type in each spot became relatively stable, indicating that a consistent positive threshold could be determined for each sub cell type (**Fig. S8**). Following the methodology of a published study^65^, we set the positive threshold for cell type A as the mean plus two times the standard deviation (“μ+2σ”) of the 130 “pseudo” estimated proportions of cell type A across all spots. If the true estimated proportion of cell type A in a spot exceeded its positive threshold, we considered cell type A to be present in that spot.

### Statistical analyses

#### Identification of NPC-associated susceptibility genes and pathways through SNP-set based genetic association analyses

Gene level - common variants based tests: we used MAGMA (version 1.07)^66^ to perform the gene-based SNP-set association analysis for common SNPs of NPC. The GWAS summary statistics of 5,503,891 common SNPs (MAF>0.01)^7,17^ was input to perform the analysis. SNPs were mapped to genes based on the gene location in NCBI 37.3^67^ with extended 35 kilobase upstream and 10 kilobase downstream window around each gene. We used the genotype of control samples in our GWAS cohort as reference population to estimate LD between variants.

Gene set level - common variants based tests: in addition to gene-based association test we performed gene set-based association test with MAGMA for common variants (version 1.07)^66^. Result of the above mentioned gene-based analysis which contained 18,063 genes’ associated P-value was used as input. A total of 13,155 gene sets were downloaded from Molecular Signature Database v2023.1.Hs^30^, including 50 HALLMARK pathways, 10,532 Gene Ontology (7,751 GO Biological Process, 1,009 GO Cellular Component and 1,772 GO Molecular Function) pathways, 186 KEGG pathways, 1,654 REACTOME pathways and 733 WikiPathways pathways. Multiple testing correction was performed using the False Discover Rate (FDR) method.

Gene level - rare variants based tests: the gene-based association analysis was performed for rare SNPs in comparisons between NPC cases with family history (labeled as “rare(FHNPC)”) vs controls and between sporadic NPC cases (labeled as “rare(SPNPC)”) vs controls, respectively. To restrict the analysis to functional rare variants, we only included SNPs with a MAF<0.01 and annotated to be functional. A SNP was defined as functional if it affected coding (non-synonymous or splice affecting); or ranked at top 5% in any of the measurements: CADD (Combined Annotation Dependent Depletion) score^68^, DANN (deleterious annotation of genetic variants using neural networks)^69^, fitCons (fitness consequence score)^70^; or listed in InterVar database (referring to clinical interpretation of missense variants)^71^. Gene-based association test was performed by an ensemble method that simultaneously tests four algorithms of SNP-set, (Sequence) Kernel Association Test (SKAT)^72^ and Burden test^73^. Details of the methods have been described in our previous study^9^. Top 10 genetic principal components and sex were jointly fitted as covariates.

Gene set level - rare variants based tests: this study conducted gene set level genetic association analysis for the aforementioned rare variants using the ensembl method and covariates aforementioned in gene level test section. Pathway data were sourced from the Molecular Signature Database v6.2^30^, with FDR correction employed for multiple testing correction of pathway association statistical results.

#### Single cell level and cell type level genetic association analysis

We used scDRS (version v1.0.1)^23^ to calculate the normalized genetic-association scores (GASs) for each cell and test the enrichment of genetic signal for each cell type for NPC. In brief, scDRS calculated normalized disease scores for individual cells based on the aggregated expression of a set of putative disease genes weighted by the significant level of gene-level association and inverse-weighted by technical noise^23^. In our study, we calculated three normalized scores for each cell: a common genetic association score (common GAS), a rare (FHNPC) genetic association score (rare(FHNPC) GAS) and a normalized rare (SPNPC) genetic association score (rare(SPNPC) GAS). In order to calculate the common or the rare GASs, summary statistics of gene-based association analyses of NPC was first used to identify putative NPC associated genes using “munge-gs” function in scDRS based on a genetic association P-value threshold of 0.005. A total of 314, 269 and 186 putative genes were used in construct the common, rare(FHNPC), rare(SPNPC) GASs, respectively (**Supplementary Tables 1**).

The following analyses were then performed for two cohorts of scRNA-seq data separately. First, the expression matrix and gene-based association statistics were provided for the ‘‘compute-score’’ function in scDRS to calculate the three GASs for each cell. To eliminate batch effects from data sources and patients, this analysis included data sources and patients as covariates. Cell type level analysis was then performed to identify the major cell types and sub cell types significantly associated with NPC (meaning significantly enriched for genetic associations) using “perform-downstream” function with default parameters. Only top 5% quantile of the GASs of a given cell type was used to test for enrichment of genetic associations for that cell types^23^. The “sumz” function in R package metap (Version 1.8)^74^ was used to meta-analyze the P-values of cell type-level result across two scRNA-seq data cohorts. Multiple testing correction for the meta-analyzed P-values was performed using the Bonferroni method. The cell types passed the threshold of P_Bonferroni_=0.05 were considered as genetically-associated cell types of NPC and labeled as “GAS^high^” (others labeled as “GAS^low^”) in downstream analyses.

#### Profiling the molecular features of cell or cell types of interest

Consistent with method described in the “Gene set-based genetic association analysis”, pathway annotations were downloaded from Molecular Signature Database v2023.1.Hs^30^ and only pathways in which more than half of the genes are detected to be expressed in the scRNA-seq data are retained for subsequent analysis. We calculated two metrics to assess the molecular features of a given cell or cell type of interest, as the following.

##### Metrics 1: up- or down-regulated pathway of a given cell type

We first calculated cell specific pathway activity scores for individual pathways using the “AddModuleScore” function with default parameters in Seurat^63^. We then compared the cell-level pathway activity score of the cells belonging to a sub cell type of interest with that of the cells belonging to other sub cell types within the same major cell type. Specifically, for a given pathway, we performed pairwise comparisons of pathway activity scores between the sub cell type of interest and other sub cell types within the same major cell type using the Wilcoxon test (two-sided). If the cell sub cell type of interest consistently exhibited significantly higher pathway activity scores (P<0.05) compared to other sub cell types in pairwise comparisons, it indicated that within that major cell type, this pathway was most active in the cell subtype of interest, labeled as “up-regulated pathway”; conversely, if the pathway activity scores were consistently lower, it indicated the weakest activity, labeled as “down-regulated pathway”. In order to assess and visualize the activity levels of the pathways of interest within cell types, we designated the median of pathway activity score of all cells within a given cell type as the pathway activity score at the cell type level.

##### Metrics 2: differentially expressed genes (DEG) and DEG-enriched pathways of a given cell type

We first identified the DEGs in a given cell type of interest relative to other sub cell types of the same major cell type. For each sub cell type of interest, we identified differentially expressed genes (DEGs) using the “FindAllMarkers” function in the R package Seurat V.4.3.0^63^. In brief, for a given sub cell type, we first performed Wilcoxon test to identify DEGs with P_Bonferroni_adjusted_ < 0.05 and log_2_fold change > 0.5 in comparisons with other sub cell types. We also limited the DEGs to only include those expressed in at least 10% more cells of sub cell types of interest compared to other sub cell types. For each given sub cell type, we then ranked all DEGs based on their log_2_fold change and retained the top 300 genes as the refined DEGs (all DEGs were refined DEGs if less than the number of DEGs were less than 300).

Subsequently, we conducted pathway enrichment analysis on the refined DEGs using the “enricher” function in the R package clusterProfiler v4.2.2^75^. Only pathways consisting of > 5 genes and < 200 genes were used in enrichment analysis. All genes expressed in the sub cell type of interest were listed in background gene set. The Bonferroni method was used for multiple testing correction for pathways per database and P_Bonferroni_adjusted_ <0.05 was considered to be significant (labeled as “sig-DEG-enriched pathway”, and the pathways with only raw P<0.05 were labeled as “DEG-enriched pathway”).

#### Gene set variation analysis (GSVA)

To contrast molecular features across sub cell types within a given major cell type, we performed gene set variation analysis (GSVA, version 1.42.0)^76^. Average expression levels of sub cell types derived from scRNA-seq Seurat object was fed into GSVA, the union of “sig-DEG-enriched pathways” of sub cell types were used as input gene sets.

#### Prediction of cell-cell interaction through ligand and receptor pairs

To explore the potential interactions between cell types, we predicted cell-cell interactions by estimating interaction probability between ligand and receptor pairs. This was realized by processing normalized expression matrix in forms of Seurat object of using CellChat (version 1.6.1) in R^77^. The analyses for the two scRNA-seq cohorts were performed separately. CellChatDB provided a list of known ligand receptor pairs (N_LR-pair_=1,792) for which we calculated the interaction probability to obtain significantly interacted LR-pairs (P_LR-interaction_ < 0.05) for any given cell type pair. The prediction of cell-cell interaction through LR was performed for every sub-cell-type-pair.

#### Identification of the key ligands and receptors contributing to the increased cell-cell interaction between cell types

We aim to identify specific ligands and receptors contributing to the interactions between the sub cell types we focused. We first extracted all significantly interacted ligand-receptor pairs between any two sub cell types (identified in “Prediction of cell-cell interaction” section). Then, we employed the one-sample Wilcoxon test to compare the interaction probabilities of the aforementioned significantly interacted ligand-receptor pairs in the sub cell type pair of interest and those of the rest of sub cell type pairs from the same major cell type categories. FDR correction was applied for the results from multiple comparisons of ligand-receptor pairs. The ligand-receptor pair with significantly higher probability in focused sub cell types (P_FDR_<0.05) in both scRNA-seq cohorts was recognized as key ligand-receptor pair contributing to the increased cell-cell interaction between the two sub cell types.

#### Comparison of cell-cell interactions between genetically stratified cell groups

The sum of the interact probability of all LR-pairs (labeled as “sum probability”) was used to represent the interaction strength for two sub cell types (a cell type pair). Only cells belonging to a major cell type containing at least one genetically-associated sub cell type (also known as “GAS^high^”) were used in the following analysis. If a cell type pair was formed by two “GAS^high^” sub cell types, we called it a “GAS^high^-GAS^high^” pair. Similarly, “GAS^low^-GAS^low^” was formed by two “GAS^low^” sub cell types and “GAS^high^-GAS^low^” was formed by one “GAS^low^” sub cell type and one “GAS^high^” sub cell type. The “non(GAS^high^-GAS^high^)” sub cell type pairs were composed of “GAS^low^-GAS^low^” and “GAS^high^-GAS^low^” sub cell type pairs. The interaction levels of all “GAS^high^-GAS^high^” pairs VS all “non(GAS^high^-GAS^high^)” pairs or VS all “GAS^low^-GAS^low^” pairs cell type pairs were compared by Wilcoxon test. Apart from comparing the overall interaction strength (sum probability), we additionally performed paired Wilcoxon test with LR-pairs matched.

To focus on the interaction of common and rare genetic associations at intercellular level, we specifically examined the sub cell type pairs consisted one cell type enriched with common genetic associations (GAS^high(common)^) and one cell type enriched with rare genetic associations (GAS^high(rare)^) by using Wilcoxon test to compare their interaction strength with that of other cell types pairs composed of the GAS^low^ counterparts within the same major cell categories. These major cell types included B, DC and Monocyte-Macrophage for common variant association and EP and iCAF for rare variant association. Such analyses were labeled as “common - rare(FHNPC)” or “common - rare(SPNPC)”. Likewise, “common - common” and “rare(FHNPC) - rare(SPNPC)” analyses were also performed.

#### Testing for the correlation between the significant level of genetic association and the strength of intercellular interactions

The correlation test was performed between the summed interaction probability of sub cell type pairs and the summed mean of GASs of the two cell types from each cell type pair. In detail, the summed interaction probability was calculated as described in “Comparison of cell-cell interactions between genetically stratified cell groups” section and was used to represent intercellular interaction strength for each sub cell type pair. The mean of all cells’ GASs within a given sub cell type was called “mean GAS”. For each sub cell type, we have a mean GAS for common variant association, a mean GAS for rare(FHNPC) variant association, a mean GAS for rare(SPNPC) variant association. Meanwhile, we selected the maximum value of rare (FHNPC) GAS and rare (SPNPC) GAS for each cell as the “rare GAS,” representing the overall rare genetic association level of the cell, and the corresponding mean GAS could be obtained. The summed mean GAS of a sub cell type pair can then be calculated by adding the mean GAS of the two sub cell types together (the mean GAS was scaled before the calculation), representing the level of genetic association of a given cell type pair. Now, the Pearson correlation can be estimated between the summed mean GAS and the summed interaction probability.

In addition to test the summed mean GAS based on common, rare(FHNPC) and rare(SPNPC) variant association separately, we also defined “max(common, rare)” and “sum(common, rare)” scores as alternative metrics to characterize the overall genetic association levels of cells. The “max(common, rare)” score represents the maximum value of common GAS and rare GAS for each cell, while the “sum(common, rare)” score represents the sum of common GAS and rare GAS for each cell. Both “max(common, rare)” and “sum(common, rare)” scores represent the global genetic association level of each cell, also referred to “Global GAS”. The summed mean GAS at the sub cell type pair level is also calculated accordingly. These analyses were performed both globally and within individual major cell type categories. Each scRNA-seq cohort was tested separately, and the consistency of the results in terms of the correlation direction (positive or negative) across two cohorts were also checked.

#### Measurements of spatial distance between cell types

Three scores, including colocalization score, normalized neighborhood score and normalized proximity score, were calculated to evaluate the space distance between any two sub cell types in spatial transcriptome data. The scores were computed in each sample independently under the following rules:

The colocalization score was computed as the Spearman correlation of two sub cell types’ proportion in each spot (derived from RCTD^26^).

The normalized neighborhood score was calculated as the following. Remembering that we had recognized whether a given sub cell types existed in a given spot or not, as described in “Cell type deconvolution of spatial transcriptome” section. Given the hexagon-shaped spot unit of spatial transcriptome data, each spot can only be surrounded by a maximum of 6 other spots. Let n_A_ donate the number of spots that contain cell type A. Let *n*_A ∪ A neighbor_ denote the total number of spots that contain cell type A as well as the spots adjacent to those containing cell type A, *n*_B ∩ (A ∪ A neighbor)_ denote the total number of spots that contain cell type B among the spots that contain cell type A as well as the spots adjacent to those containing cell type A. In other words, *n*_B ∩ (A ∪ A neighbor)_ is the total number of B neighbors of A. We could now calculate the raw neighborhood score for the cell type B around cell type A (cell type A as the core, labeled as “A(core)-B”) as:

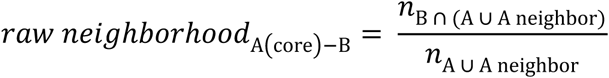

Since the raw neighborhood_A(core)-B_ will be influenced by overall distribution of cell type B, we further derived the normalized neighborhood score:

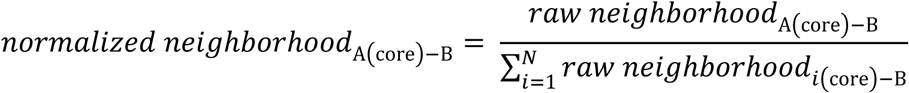

Where N is the total number of sub cell types annotated to the given spatial transcriptomic sample.

The normalized proximity score was calculated by describing the address of all spots in a given sample to a two dimension XY coordinate system, where (*x*_A1_, *y*_A1_), (*x*_A2_, *y*_A2_),… (*x*_A*n*_A__, *y*_A*n*_A__) donates the address of *n*_A_ spots containing cell type A (similarly for cell type B). The unit of the coordinates in each axis is micrometre (μm). We first take a spot with a cell type A as the core, assuming its coordinates are (*x*_A*i*_, *y*_A*i*_). Then, we select any spot with a cell type B, assuming its coordinates are (*x*_B*i*_, *y*_B*i*_). The distance from this spot with cell type B to the spot with cell type A can be calculated as follows:

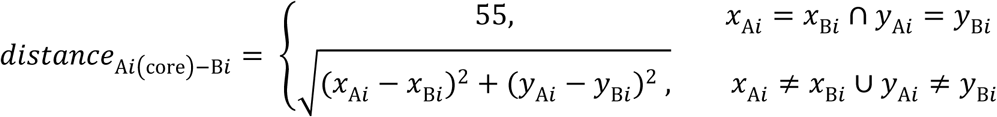

When sub cell type A and B colocalize on the same spot, the *distance*_*Ai*(*core*)*Bi*_ is assigned a constant value of 55 because the diameter of a spot on the Visium spatial transcriptome data is 55μm.

When we take a spot with a cell type A as the core, in fact, the distance between all spots with cell type B and the focused spot with cell type A can be calculated using the formula mentioned above. We select the spot with cell type B that is closest to the spot with cell type A and calculate its distance. This distance represents the distance between this spot with cell type A as the core and the cell type B, which is:

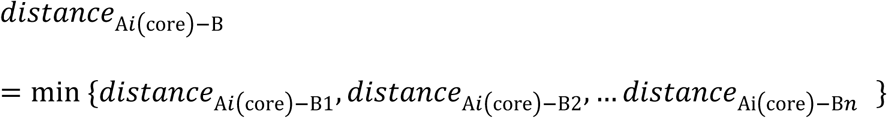

For the given sample, there are a total of *n_A_* spots with cell type A. When each of these spots is considered as the core, the distances to cell type B provide overall distance information for the cell type pair A(core)-B. Now, for the cell type pair A(core)-B, their original proximity score between them can be calculated as follows:

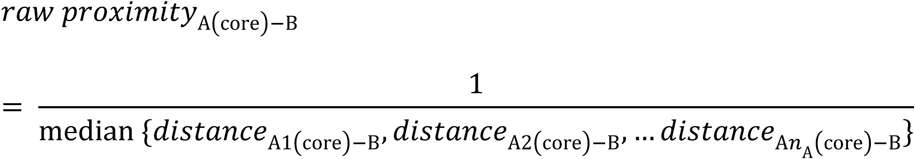

Since the raw proximity_A(*core*)-B_ may also be influenced by the overall distribution of cell type B, similar to the normalization of the neighborhood scores, we use a normalized proximity score in our downstream analysis as follows:

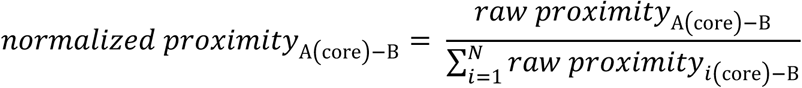

#### Significant tests for the spatial association between two cell types

In order to test whether the observed level of colocalization or neighborhood between any given cell type pair is stronger than what would be expected by chance given the overall distribution of the two cell types, we performed Fisher’s exact test (one sided) to test the significant spatial association between the two cell types. Specifically, 1) colocalization enrichment: we tested whether cell type A located spots significantly overlapped with spots located cell type B; 2) neighborhood enrichment: we tested whether cell type A located spots and spots surrounded significantly overlapped with spots located cell type B and spots surrounded. The tests were performed in each ST sample independently, and we used the “sumz” function in R package metap^74^ to meta-analyze P-value of all samples.

#### Comparison of spatial distance between genetically stratified cell types

We used Wilcoxon test to compare the colocalization, normalized neighborhood and normalized proximity scores of “GAS^high^-GAS^high^” with “GAS^high^-GAS^low^” and with “GAS^low^-GAS^low^” sub cell type pairs. Specific comparisons were additionally performed for “common - rare”, “common - rare(FHNPC)”, “common – rare(SPNPC)” and “rare(FHNPC) - rare(SPNPC)” pairs.

#### Prioritizing cell type pairs by integrating cell-cell interaction and spatial distance information

To identify genetically-associated cell type pairs that display both strong intercellular interactions and close spatial distance, for each sub cell type pair, we calculated and ranked the median values of the scaled (mean of 0 and standard deviation of 1) following four metrices: summed interaction probability (interaction strength), colocalization score, normalized neighborhood score, and normalized proximity score. The sub cell type pair with highest median value were identified as the prioritized pair.

#### Identification of direct genetic contributions to cell-cell interactions – encoding ligands and receptors

NPC-associated susceptibility genes could directly contribute to intercellular interactions by encoding ligand/receptor genes. We used Fisher’s exact test to assess whether there is a significant overlap of the ligand (L)/receptor (R) genes encoding significantly interacted LR-pairs for cell-cell interactions with NPC-associated susceptibility genes. Here, the NPC-associated susceptibility genes were identified using P-value<0.05 as the threshold in GWAS or EWAS-based gene-level SNP-set association tests. The significantly interacted LR-pairs were also identified using the of threshold P-value<0.05 in cell-cell interaction analysis by CellChat, as described in the “Prediction of cell-cell interaction (through LR-pairs)” section.

#### Identification of indirect genetic contributions to cell-cell interactions – LR associated pathways

NPC’s genetic risk genes could participate the intercellular interactions indirectly through involved in pathways interplaying with LR signaling. For this, we first tested the Spearman correlation between the expression levels of the key LR genes and the GASs (genetic association score) of cells in the target cell type. The spearman correction tests were carried out in two scRNA-seq cohorts (“C1” and “C2”) respectively. After that, we conducted the Meta-analysis of corrections across two cohorts using function “metacor” in R package meta (Version 6.5.0)^78^ with default parameters. The Fisher’s z transformation was used to estimate the meta-analyzed correlation coefficient (labeled as “meta-cor”) using random effects model. If a significant correlation was detected between any key LR gene and GAS, that means at least some of the putative associated genes used to derive GAS either directly encode the key LR genes (implicating direct contribution) or participate/influence the pathways associated with that LR gene (implicating indirect contribution). We have tested the former condition in previous section and for the later condition, we performed Spearman correlation tests between LR gene expression levels and pathway activity scores in a cell-type-specific manner, which allowed us to identify pathways significantly correlated (R>0.1, P_FDR_<0.05) with the expression of the indirect-effect-targeted LR genes. We labeled such identified pathways as “ligand/receptor-associated pathways, and performed a series of analyses on them.

First, as the GASs represented the NPC-associated genetic loading of a given cell, to test whether the ligand/receptor-associated pathways were enriched with genetic association signals, we calculated the correlation of the ligand/receptor-associated pathway activity scores with GASs, and compared the correlation coefficient to that of the non-ligand/receptor-associated pathways using Wilcoxon test.

Meanwhile, beyond the GASs themselves, genetic associations can be enriched in pathways associated with the GASs. A significant overlap between GAS-associated pathways and ligand/receptor-associated pathways would also indicate a high genetic loading of ligand/receptor-associated pathways. For this, we identified GAS-associated pathways through Spearman correlation tests between GAS and pathway activity scores. Significantly correlated pathways with a R>0.1and a P_FDR_<0.05 were identified as GAS-associated pathways (GAS-assoc pathways) for a given cell type. We then tested if the GAS-assoc pathways have a significant overlap with the ligand/receptor-associated pathways using Fisher’s exact test.

Additionally, we examined whether ligand/receptor-associated pathways significantly overlap with the NPC-associated susceptibility pathways identified by gene-set-level association tests (genetic-assoc pathways, identified in “SNP-set based genetic association analyses” section). The genetic-assoc pathways were identified based on a significance threshold of P<0.005. Fisher’s exact test was applied to test the significance of the overlap of those pathways.

Furthermore, to elucidate whether ligand/receptor-associated pathways participate in the core functions of the target sub cell type, we tested whether there is a significant overlap between functionally featuring pathways for the target sub cell type and the ligand/receptor-associated pathways. The functionally featuring pathways included up-regulated and DEG-enriched pathways which we identified in the “Profiling the molecular features of cell or cell types of interest” section. Fisher’s exact test was applied to test the significance of the overlap. FDR correction was used for multiple testing correction of P-values in the above tests. The collection of pathway information and the generation of pathway activity scores used in all aforementioned analyses have been described in the “Profiling the molecular features of cell or cell types of interest” section.

#### Survival analysis based on infiltration levels of specific sub cell types

To calculate enrichment scores for the cell types of interest based on marker genes specific for those cell types in bulk RNA-seq data, Single-sample Gene Set Enrichment Analysis (ssGSEA) was conducted using the “run_ssGSEA2” function with default parameters in R package ssGSEA2^39^. Specifically, the sub-cell-type-specific marker genes were identified through the “FindAllMarkers” function of Seurat^63^, which detects specifically overexpressed marker genes for the corresponding sub cell type. A gene was identified as a candidate marker gene if it satisfies all of the following three conditions: (1) Wilcoxon test showed its expression significantly upregulated (P_Bonferroni_adjusted_ < 0.05) with log2fold change > 1 in the sub cell type compared to all other cells and (2) the difference between the percentage of expressed cells in the target sub cell type (PCT_target_) and the percentage of expressed cells in the rest of all cells (PCT_rest_) is higher than 0.3 (PCT_target_ – PCT_rest_ > 0.3) or the ratio of PCT_target_ and PCT_rest_ was higher than 3 (PCT_target_ / PCT_rest_ > 3); (3) its expression levels in the target sub-cell type were significantly higher (P<0.05) than those in each of the other sub-cell types using Wilcoxon tests in a stepwise manner. The genes selected through the above steps were used as marker genes for target sub cell type and were used in ssGSEA analysis. The resulted enrichment scores represented the infiltration level of the cell type of interest in the tumor tissues^39^. Samples with an enrichment score (infiltration level) beyond the range of mean plus or minus three times the standard deviation (μ±3σ) were identified as outliers and removed in subsequent survival analysis.

Cox regression survival analyses were then performed using the “coxph” function in R package survival (Version 3.5.5)^79^ to test for the association between the infiltration level of the cell type of interest and the prognosis of NPC patients (measured as disease free survival, DFS). Gender and batch were fitted as covariates. In addition to testing the infiltration levels of each target sub cell types separately, we also tested the effects from the ratio of the infiltration levels of two target sub cell types, the product of the infiltration level of two target sub cell types, as well as simultaneously fitting infiltration levels of the two target sub cell types in Cox regression models. To compare the performance of the models with different predictors, we used “coxr2” function in R package CoxR2 (Version 1.0)^80^ to calculate the R^2^ of each model. The likelihood ratio test was conducted with “lrtest” function in R package lmtest (Version 0.9.40)^81^ to compare the goodness of fit between two nested models.

Visualization of the differentiated survival under stratified infiltration levels was realized by Kaplan-Meier curve with the “survfit” function in R package survival^79^. The P-value used to test the difference between two groups was derived from log-rank test, besides, we fitted a stratified Cox proportional-hazards model to estimate the hazard ratio (HR) and its 95% confidence interval (CI).

## Data Availability

The genotype data generated by WES have been deposited in the Genome Variation Map repository (https://ngdc.cncb.ac.cn/gvm/) of National Genomics Data Center (NGDC) under controlled access due to data privacy laws related to patient consent for data sharing with accession number GVM000580. The NPC bulk RNA-seq data are deposited at the OMIX repository (https://ngdc.cncb.ac.cn/omix/) of the NGDC (OMIX004586) and the GEO database (GSE102349). The NPC single-cell RNA-seq data are available in the GEO database (GSE162025, GSE150825, GSE150430) and GSA database under accession code HRA000087. The spatial transcriptomic data are available in GEO database (GSE206245).

https://ngdc.cncb.ac.cn/gvm/getProjectDetail?project=GVM000580

https://ngdc.cncb.ac.cn/omix/release/OMIX004586

https://www.ncbi.nlm.nih.gov/geo/query/acc.cgi?acc=GSE102349

https://www.ncbi.nlm.nih.gov/geo/query/acc.cgi?acc=GSE162025

https://www.ncbi.nlm.nih.gov/geo/query/acc.cgi?acc=GSE150825

https://www.ncbi.nlm.nih.gov/geo/query/acc.cgi?acc=GSE150430

https://www.ncbi.nlm.nih.gov/geo/query/acc.cgi?acc=GSE206245

https://ngdc.cncb.ac.cn/gsa-human/browse/HRA000087

## Study approval

The study was approved by the Sun Yat-sen University Cancer Center (SYSUCC) Ethics Committee in Guangzhou, China (reference no. SL-B2021-032-03).

## Acknowledgement

We acknowledge supports from the National Natural Science Foundation of China (NSFC; 82261160657), the National Key R&D Program of China (NKRDPC; 2022YFC3400901), the Chang Jiang Scholars Program (J.X.B.).

## References

1. Cai, Y. et al. An atlas of genetic effects on cellular composition of the tumor microenvironment. Nat Immunol 25, 1959–1975 (2024).

2. Baghban, R. et al. Tumor microenvironment complexity and therapeutic implications at a glance. Cell Commun Signal 18, 59 (2020).

3. Lin, Q. et al. Single-cell analysis reveals the multiple patterns of immune escape in the nasopharyngeal carcinoma microenvironment. Clinical & Translational Med 13, e1315 (2023).

4. Liu, Y. et al. Single-cell and spatial transcriptome analyses reveal tertiary lymphoid structures linked to tumour progression and immunotherapy response in nasopharyngeal carcinoma. Nat Commun 15, 7713 (2024).

5. Sud, A., Kinnersley, B. & Houlston, R. S. Genome-wide association studies of cancer: current insights and future perspectives. Nat Rev Cancer 17, 692–704 (2017).

6. Wilcox, N. et al. Exome sequencing identifies breast cancer susceptibility genes and defines the contribution of coding variants to breast cancer risk. Nat Genet 55, 1435–1439 (2023).

7. Bei, J.-X. et al. A genome-wide association study of nasopharyngeal carcinoma identifies three new susceptibility loci. Nat Genet 42, 599–603 (2010).

8. He, Y.-Q. et al. A polygenic risk score for nasopharyngeal carcinoma shows potential for risk stratification and personalized screening. Nat Commun 13, 1966 (2022).

9. Zeng, Y. et al. Whole-exome sequencing association study reveals genetic effects on tumor microenvironment components in nasopharyngeal carcinoma. J Clin Invest 135, e182768 (2025).

10. Wang, C. et al. Analyses of rare predisposing variants of lung cancer in 6,004 whole genomes in Chinese. Cancer Cell 40, 1223–1239.e6 (2022).

11. Mandal, R. et al. Genetic diversity of tumors with mismatch repair deficiency influences anti–PD-1 immunotherapy response. Science 364, 485–491 (2019).

12. Watanabe, K. et al. A global overview of pleiotropy and genetic architecture in complex traits. Nat Genet 51, 1339–1348 (2019).

13. Pagadala, M. Germline modifiers of the tumor immune microenvironment implicate drivers of cancer risk and immunotherapy response. Nature Communications (2023).

14. Chu, X. et al. Integrative single-cell analysis of human colorectal cancer reveals patient stratification with distinct immune evasion mechanisms. Nat Cancer 5, 1409– 1426 (2024).

15. Law, P. J. et al. Systematic prioritization of functional variants and effector genes underlying colorectal cancer risk. Nat Genet 56, 2104–2111 (2024).

16. Ding, B. et al. Comprehensive single-cell analysis reveals heterogeneity of fibroblast subpopulations in ovarian cancer tissue microenvironment. Heliyon 10, e27873 (2024).

17. Cui, Q. et al. An extended genome-wide association study identifies novel susceptibility loci for nasopharyngeal carcinoma. Hum. Mol. Genet. 25, 3626–3634 (2016).

18. Liu, Y. et al. Tumour heterogeneity and intercellular networks of nasopharyngeal carcinoma at single cell resolution. Nat Commun 12, 741 (2021).

19. Gong, L. et al. Comprehensive single-cell sequencing reveals the stromal dynamics and tumor-specific characteristics in the microenvironment of nasopharyngeal carcinoma. Nat Commun 12, 1540 (2021).

20. Chen, Y.-P. et al. Single-cell transcriptomics reveals regulators underlying immune cell diversity and immune subtypes associated with prognosis in nasopharyngeal carcinoma. Cell Res 30, 1024–1042 (2020).

21. Jin, S. et al. Single-cell transcriptomic analysis defines the interplay between tumor cells, viral infection, and the microenvironment in nasopharyngeal carcinoma. Cell Res 30, 950–965 (2020).

22. Zhao, J. et al. Single cell RNA-seq reveals the landscape of tumor and infiltrating immune cells in nasopharyngeal carcinoma. Cancer Letters 477, 131–143 (2020).

23. Zhang, M. J. et al. Polygenic enrichment distinguishes disease associations of individual cells in single-cell RNA-seq data. Nat Genet https://doi.org/10.1038/s41588-022-01167-z (2022) doi:10.1038/s41588-022-01167-z.

24. Su, Z. Y., Siak, P. Y., Leong, C.-O. & Cheah, S.-C. Nasopharyngeal Carcinoma and Its Microenvironment: Past, Current, and Future Perspectives. Front. Oncol. 12, 840467 (2022).

25. Roma-Rodrigues, C., Mendes, R., Baptista, P. & Fernandes, A. Targeting Tumor Microenvironment for Cancer Therapy. IJMS 20, 840 (2019).

26. Cable, D. M. et al. Robust decomposition of cell type mixtures in spatial transcriptomics. Nat Biotechnol 40, 517–526 (2022).

27. Cai, M.-B. et al. Expression of Human Leukocyte Antigen G is associated with Prognosis in Nasopharyngeal Carcinoma. Int. J. Biol. Sci. 8, 891–900 (2012).

28. Yang, K., Halima, A. & Chan, T. A. Antigen presentation in cancer — mechanisms and clinical implications for immunotherapy. Nat Rev Clin Oncol 20, 604–623 (2023).

29. MacNabb, B. W. et al. Dendritic cells can prime anti-tumor CD8+ T cell responses through major histocompatibility complex cross-dressing. Immunity 55, 982–997.e8 (2022).

30. Subramanian, A. et al. Gene set enrichment analysis: A knowledge-based approach for interpreting genome-wide expression profiles. Proc. Natl. Acad. Sci. U.S.A. 102, 15545–15550 (2005).

31. Van Rhijn, I. Do antigen-presenting CD1a, CD1b, CD1c, and CD1d molecules bind different self-lipids? Trends in Immunology 44, 757–759 (2023).

32. Lenormand, C., et al. *HLA-DQA2* and *HLA-DQB2* Genes Are Specifically Expressed in Human Langerhans Cells and Encode a New HLA Class II Molecule. The Journal of Immunology 188, 3903–3911 (2012).

33. Uto, T. et al. Clec4A4 Acts as a Negative Immune Checkpoint Regulator to Suppress Antitumor Immunity. Cancer Immunology Research 11, 1266–1279 (2023).

34. Yasuda, K., Nakanishi, K. & Tsutsui, H. Interleukin-18 in Health and Disease. IJMS 20, 649 (2019).

35. Shen, D. et al. Melatonin inhibits bladder tumorigenesis by suppressing PPARγ/ENO1-mediated glycolysis. Cell Death Dis 14, 246 (2023).

36. Sharma, D., Singh, M. & Rani, R. Role of LDH in tumor glycolysis: Regulation of LDHA by small molecules for cancer therapeutics. Seminars in Cancer Biology 87, 184–195 (2022).

37. Fernandez-Checa, J. C., Torres, S. & Garcia-Ruiz, C. HILPDA, a new player in NASH-driven HCC, links hypoxia signaling with ceramide synthesis. Journal of Hepatology 79, 269–272 (2023).

38. Zhang, J. & Ney, P. A. Role of BNIP3 and NIX in cell death, autophagy, and mitophagy. Cell Death Differ 16, 939–946 (2009).

39. Krug, K. et al. A Curated Resource for Phosphosite-specific Signature Analysis. Molecular & Cellular Proteomics 18, 576–593 (2019).

40. Gibson, G. Rare and common variants: twenty arguments. Nat Rev Genet 13, 135–145 (2012).

41. Tufail, M., Jiang, C.-H. & Li, N. Immune evasion in cancer: mechanisms and cutting-edge therapeutic approaches. Sig Transduct Target Ther 10, 227 (2025).

42. Cao, S.-M., Chen, S.-H., Qian, C.-N., Liu, Q. & Xia, Y.-F. Familial nasopharyngeal carcinomas possess distinguished clinical characteristics in southern China. Chin J Cancer Res 26, 543–549 (2014).

43. OuYang, P.-Y. et al. Prognostic impact of family history in southern Chinese patients with undifferentiated nasopharyngeal carcinoma. Br J Cancer 109, 788–794 (2013).

44. Schork, N. J., Murray, S. S., Frazer, K. A. & Topol, E. J. Common vs. rare allele hypotheses for complex diseases. Current Opinion in Genetics & Development 19, 212–219 (2009).

45. Ang, Y.-S. et al. Disease Model of GATA4 Mutation Reveals Transcription Factor Cooperativity in Human Cardiogenesis. Cell 167, 1734–1749.e22 (2016).

46. Singhal, P. et al. Evidence of epistasis in regions of long-range linkage disequilibrium across five complex diseases in the UK Biobank and eMERGE datasets. The American Journal of Human Genetics 110, 575–591 (2023).

47. Bos, I. R. & Burkhardt, A. Epithelial and interepithelial mitoses of the oral mucosa: light and electron microscopic study in mice after exposure to different antigens. J Invest Dermatol 76, 63–67 (1981).

48. Sparber, F. Langerhans cells: an update. J Deutsche Derma Gesell 12, 1107–1111 (2014).

49. Herfs, M. et al. Transforming Growth Factor-β1-Mediated Slug and Snail Transcription Factor Up-Regulation Reduces the Density of Langerhans Cells in Epithelial Metaplasia by Affecting E-Cadherin Expression. The American Journal of Pathology 172, 1391–1402 (2008).

50. Hovav, A. & Wilensky, A. The role of the epithelial sentinels, Langerhans cells and ΓΔT cells, in oral squamous cell carcinoma. Periodontology 2000 prd.12544 (2024) doi:10.1111/prd.12544.

51. Gabrilovich, D. et al. Vascular endothelial growth factor inhibits the development of dendritic cells and dramatically affects the differentiation of multiple hematopoietic lineages in vivo. Blood 92, 4150–4166 (1998).

52. Harada, A. et al. Hypoxia-induced Wnt5a-secreting fibroblasts promote colon cancer progression. Nat Commun 16, 3653 (2025).

53. Benoot, T., Piccioni, E., De Ridder, K. & Goyvaerts, C. TNFα and Immune Checkpoint Inhibition: Friend or Foe for Lung Cancer? IJMS 22, 8691 (2021).

54. Zhao, H. et al. Inflammation and tumor progression: signaling pathways and targeted intervention. Sig Transduct Target Ther 6, 263 (2021).

55. Wijnhoven, B. P. L., Dinjens, W. N. M. & Pignatelli, M. E-cadherin—catenin cell—cell adhesion complex and human cancer. Journal of British Surgery 87, 992– 1005 (2000).

56. Ma, C. X., Jia, T. C., Li, X. R., Zhand, Z. F. & Yiao, C. B. Langerhans cells in nasopharyngeal carcinoma in relation to prognosis. In Vivo 9, 225–229 (1995).

57. Hilly, O. et al. CD1a-positive dendritic cell density predicts disease-free survival in papillary thyroid carcinoma. Pathology - Research and Practice 211, 652–656 (2015).

58. Wang, X. et al. High expression of HILPDA is an adverse prognostic prognostic factor in hepatocellular carcinoma. Medicine 102, e33145 (2023).

59. Povero, D. et al. HILPDA promotes NASH-driven HCC development by restraining intracellular fatty acid flux in hypoxia. Journal of Hepatology 79, 378–393 (2023).

60. Liu, H. et al. Nasopharyngeal carcinoma: current views on the tumor microenvironment’s impact on drug resistance and clinical outcomes. Mol Cancer 23, 20 (2024).

61. He, F. et al. Targeting the LMP1-ALIX axis in EBV+ nasopharyngeal carcinoma inhibits immunosuppressive small extracellular vesicle secretion and boosts anti-tumor immunity. Cancer Commun (Lond) 44, 1391–1413 (2024).

62. Korsunsky, I. et al. Fast, sensitive and accurate integration of single-cell data with Harmony. Nat Methods 16, 1289–1296 (2019).

63. Hao, Y. et al. Integrated analysis of multimodal single-cell data. Cell 184, 3573–3587.e29 (2021).

64. Zhang, L. et al. Genomic Analysis of Nasopharyngeal Carcinoma Reveals TME-Based Subtypes. Molecular Cancer Research 15, 1722–1732 (2017).

65. Barkley, D. et al. Cancer cell states recur across tumor types and form specific interactions with the tumor microenvironment. Nat Genet 54, 1192–1201 (2022).

66. de Leeuw, C. A., Mooij, J. M., Heskes, T. & Posthuma, D. MAGMA: Generalized Gene-Set Analysis of GWAS Data. PLoS Comput Biol 11, e1004219 (2015).

67. Sayers, E. W. et al. Database resources of the National Center for Biotechnology Information. Nucleic Acids Research 49, D10–D17 (2021).

68. Rentzsch, P., Witten, D., Cooper, G. M., Shendure, J. & Kircher, M. CADD: predicting the deleteriousness of variants throughout the human genome. Nucleic Acids Research 47, D886–D894 (2019).

69. Quang, D., Chen, Y. & Xie, X. DANN: a deep learning approach for annotating the pathogenicity of genetic variants. Bioinformatics 31, 761–763 (2015).

70. Gulko, B., Hubisz, M. J., Gronau, I. & Siepel, A. A method for calculating probabilities of fitness consequences for point mutations across the human genome. Nat Genet 47, 276–283 (2015).

71. Li, Q. & Wang, K. InterVar: Clinical Interpretation of Genetic Variants by the 2015 ACMG-AMP Guidelines. The American Journal of Human Genetics 100, 267– 280 (2017).

72. Wu, M. C. et al. Rare-variant association testing for sequencing data with the sequence kernel association test. Am J Hum Genet 89, 82–93 (2011).

73. Ionita-Laza, I., Lee, S., Makarov, V., Buxbaum, J. D. & Lin, X. Family-based association tests for sequence data, and comparisons with population-based association tests. Eur J Hum Genet 21, 1158–1162 (2013).

74. Dewey, M. metap: Meta-Analysis of Significance Values. https://CRAN.R-project.org/package=metap (2022).

75. Wu, T. et al. clusterProfiler 4.0: A universal enrichment tool for interpreting omics data. The Innovation 2, 100141 (2021).

76. Hänzelmann, S., Castelo, R. & Guinney, J. GSVA: gene set variation analysis for microarray and RNA-Seq data. BMC Bioinformatics 14, 7 (2013).

77. Jin, S. et al. Inference and analysis of cell-cell communication using CellChat. Nat Commun 12, 1088 (2021).

78. Balduzzi, S., Rücker, G. & Schwarzer, G. How to perform a meta-analysis with R: a practical tutorial. Evid Based Mental Health 22, 153–160 (2019).

79. Therneau, T. A package for survival analysis in R. https://CRAN.R-project.org/package=survival (2023).

80. Hyeri You, Ronghui Xu. CoxR2: R-Squared Measure Based on Partial LR Statistic, for the Cox PH Regression Model. 1.0 10.32614/CRAN.package.CoxR2 (2020).

81. Zeileis, A. & Hothorn, T. Diagnostic Checking in Regression Relationships. R News 2, 7–10 (2002).

