## Supplementary material for "Germline genetic risk converges through intercellular crosstalk in tumor microenvironment": Supplymentary Figure

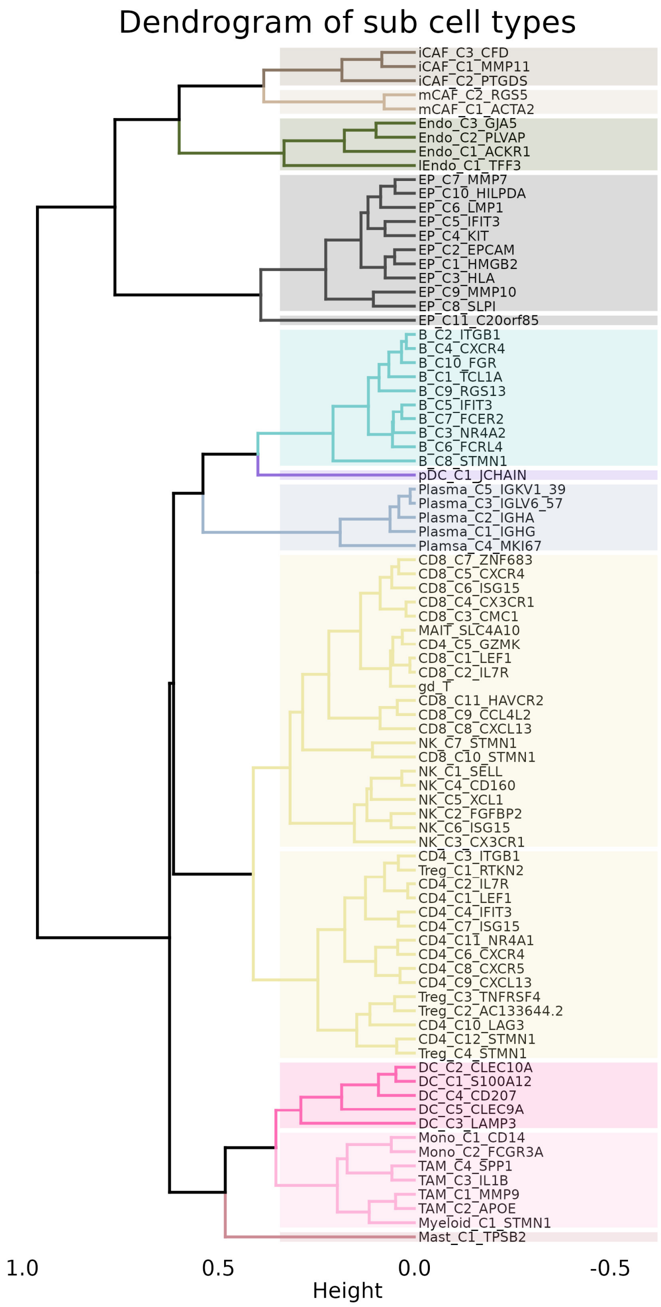


**Figure S1. Hierarchical clustering of all sub cell types in scRNA-seq data of NPC tumor samples.** All 85 sub cell types identified from NPC scRNA-seq data were hierarchically clustered to 11 clusters as major cell types. From top to bottom, the major cell types are as follows: Dark brown: Inflammatory cancer-associated fibroblasts (iCAF) cells; Light brown: Myofibroblasts cancer-associated fibroblast (mCAF) cells; Green: Endothelial cells; Gray: Epithelial cells (EP); Sky blue: B cells; Purple: Plasmacytoid dendritic cells (pDC); Dark blue: Plasma cells; Yellow: NK/T cells; Deep pink: Dendritic cells (DC); Light pink: Monocyte-Macrophage cells; Deep red: Mast cells.

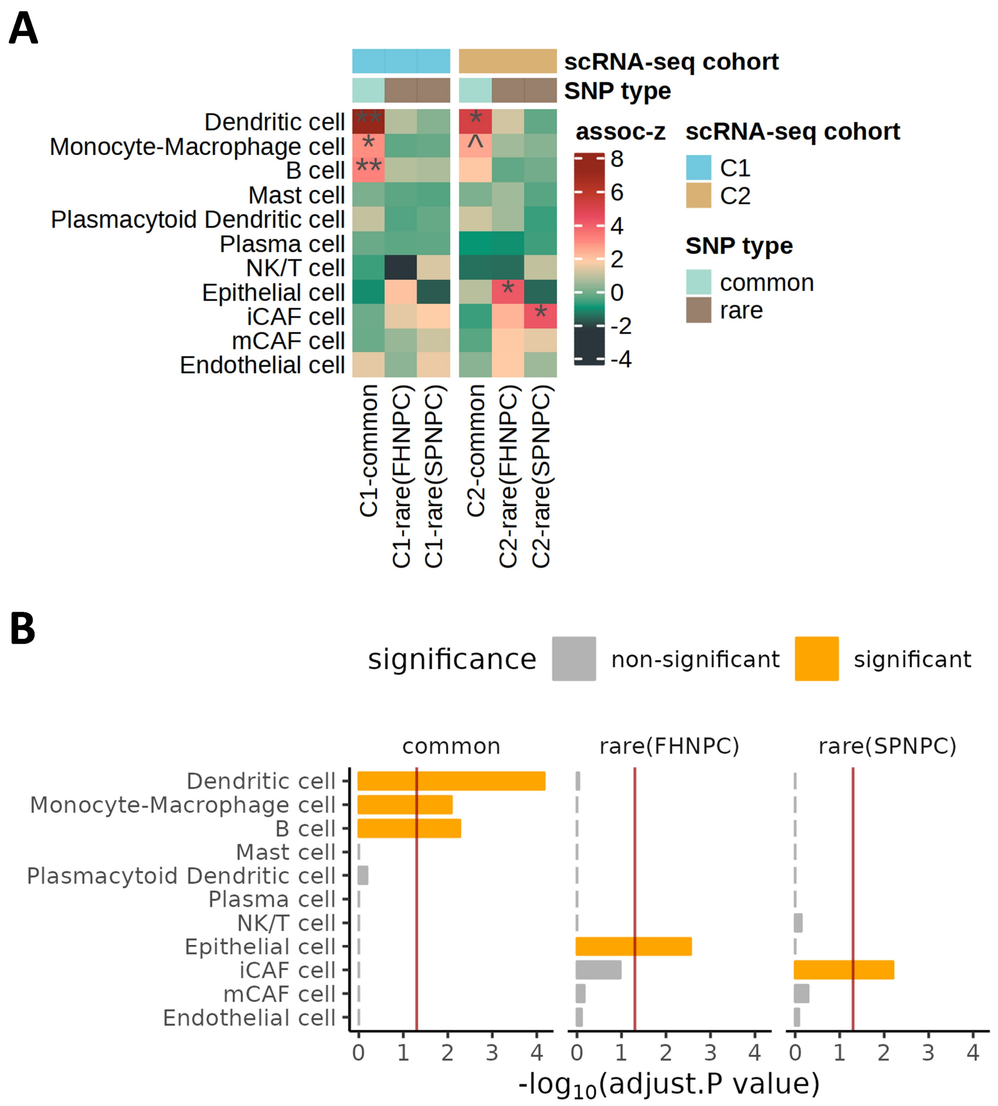


**Figure S2. Genetically-associated major cell types in NPC tumors. A.** Enrichment level of the expression of putative NPC-associated genes in 11 major cell types derived using the scDRS Methods in two scRNA-seq cohorts. The set of putative NPC-associated genes was defined and tested for common and rare genetics separately. assoc-z: representing the enrichment level for common/rare genetic association in each major cell type; significance level: ^ refers to $p<0.05$, * refers to adjust $p<0.05$, ** refers to adjust $p<0.01$. The P-values were adjusted using the FDR method. **B.** Meta-analysis for the enrichment results across two scRNA-seq cohorts' scDRS in all major cell types. Red line: adjust $p=0.05$. The P-values were adjusted using the Bonferroni method.

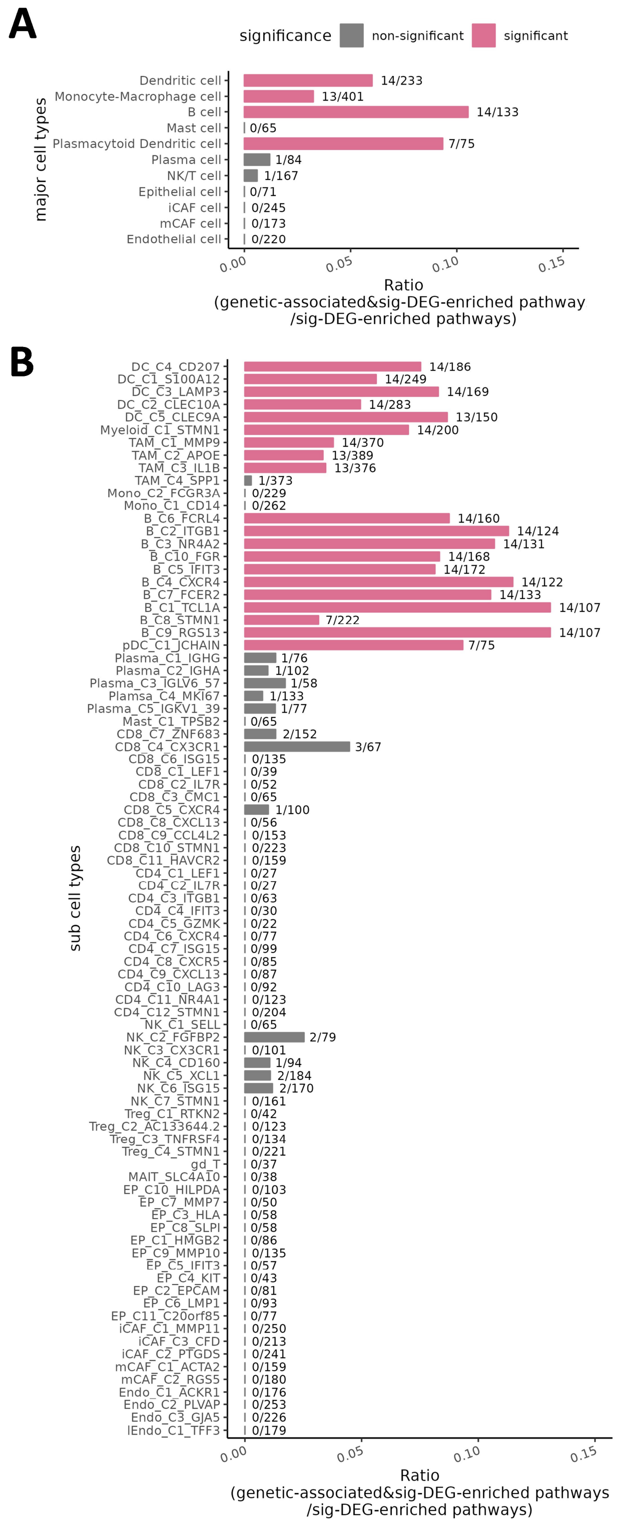


**Figure S3. Pathway overlap between genetically-associated pathways and DEG-enriched pathways of cell types.** Barplot showed the proportion of sig-DEG-enriched pathways in each of the 11 major cell types (**A**) or each of the 85 sub cell types (**B**) overlapping with genetically-associated pathways ($P_{association\text{-}FDR}$<0.05). The Fisher's exact test was used to test the significance of the overlap, $P_{FDR}$<0.1 (multiple testing correction was performed using FDR method) was considered significant and highlighted in pink.

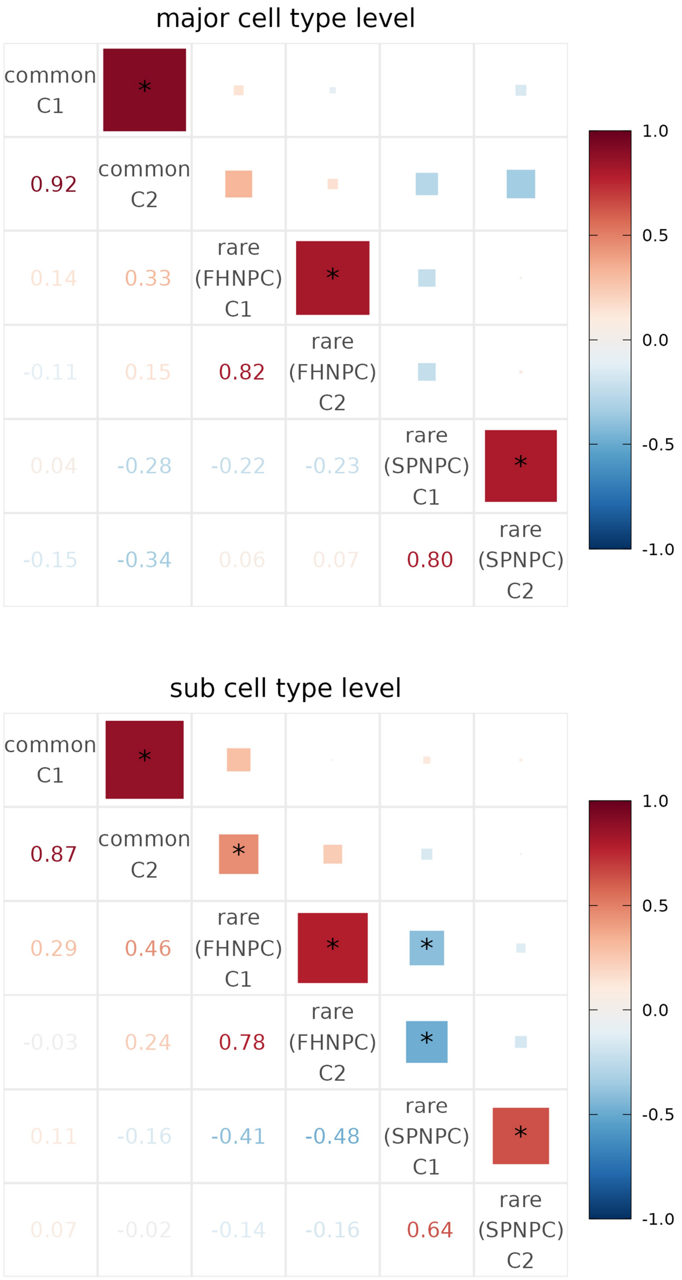


**Figure S4. The correlation of enrichment level of genetic associations in major and sub cell types between scRNA-seq cohort 1 (C1) and 2 (C2).** The test statistics, assoc-z, representing the enrichment level for common/rare genetic association in major cell types or sub cell types, was derived from scDRS method and used to calculate the correlation. The colour and size of blocks showed the coefficient of Spearman correlations. Multiple testing correction was performed using Bonferroni method (*: adjust $p<0.05$).

common

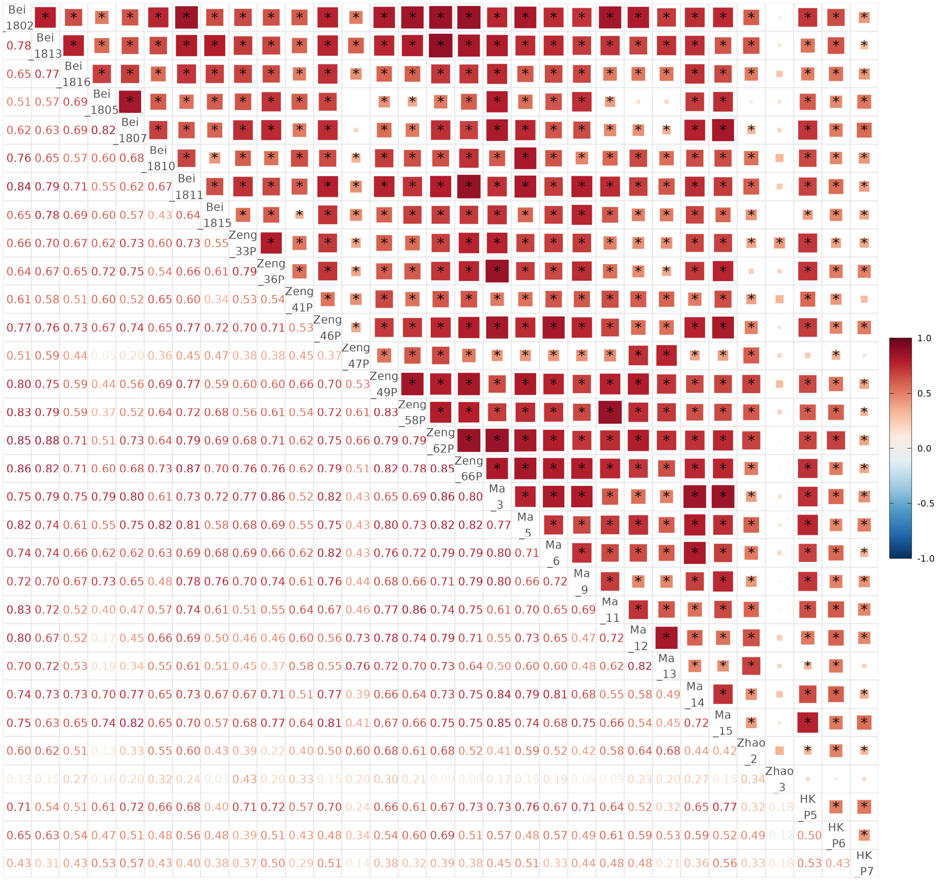


rare(FHNPC)

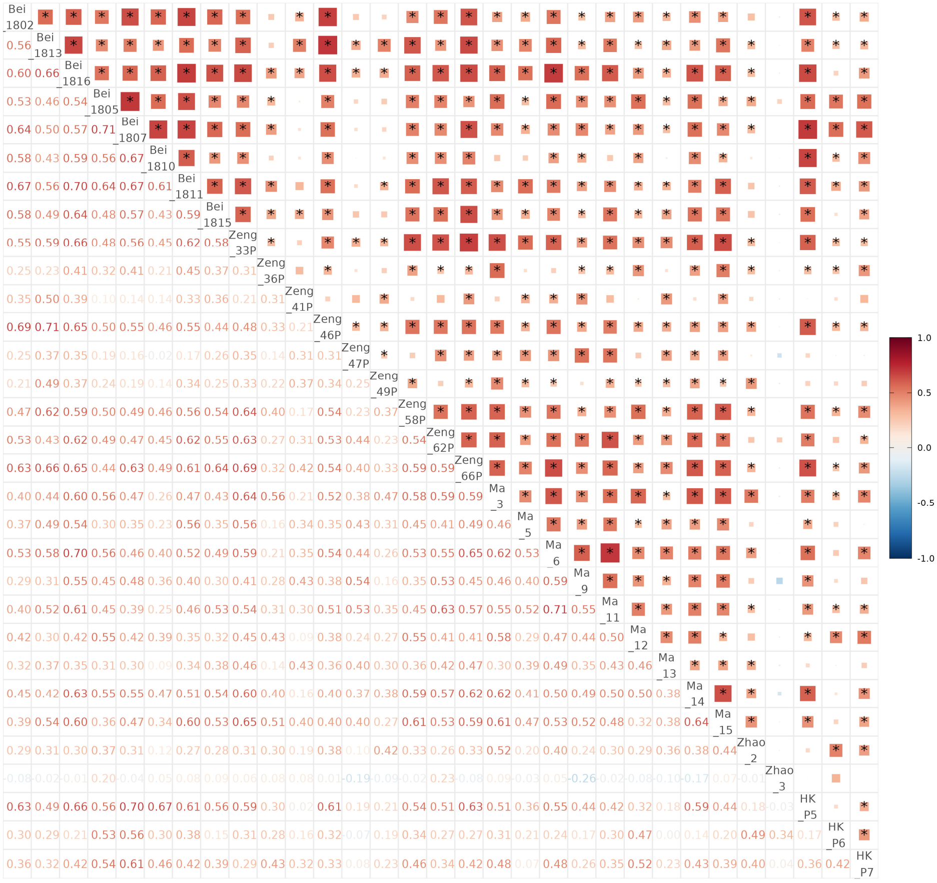

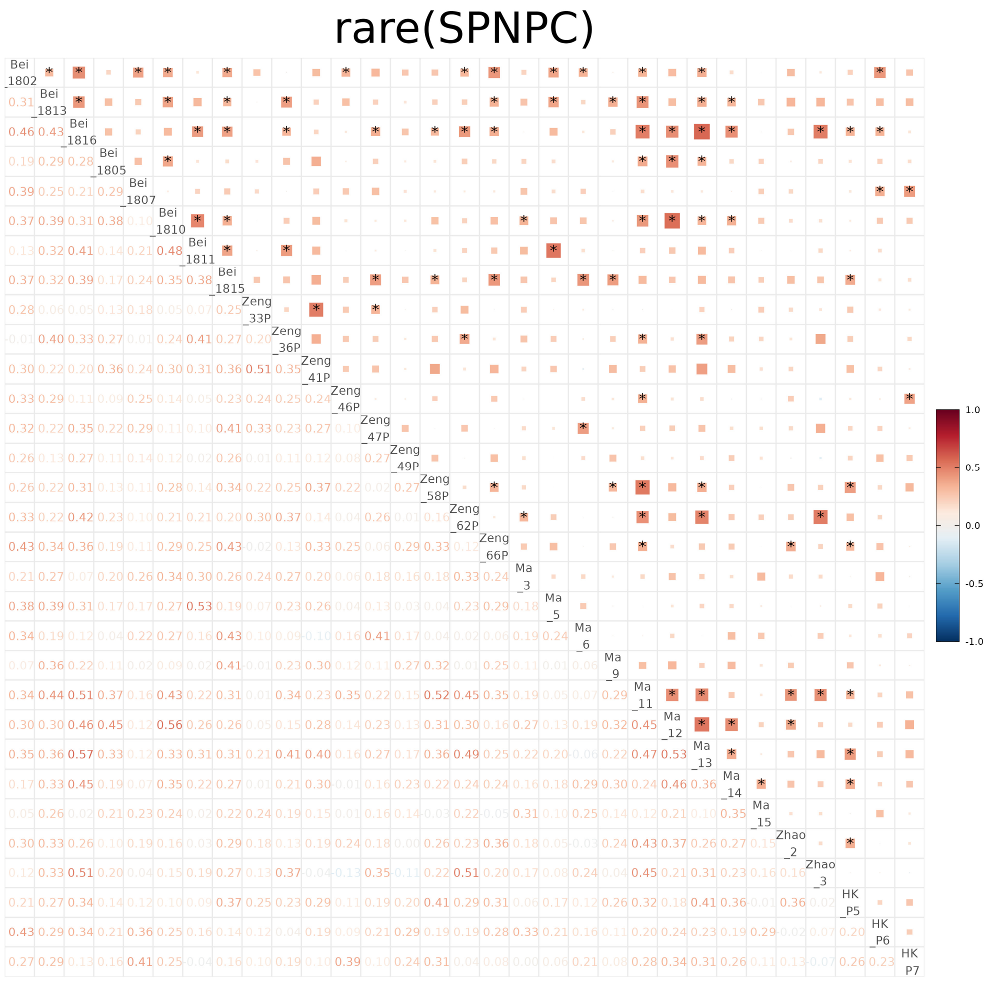

**Figure S5. The correlation of enrichment level of genetic associations in sub cell types between 31 scRNA-seq samples.** Each element represented the correlation of enrichment levels between two samples. The test statistics, assoc-z, representing the enrichment level for common/rare genetic association in sub cell types, was derived from scDRS method and used to calculate the correlation. The colour and size of blocks showed the coefficient of Spearman correlations. Multiple testing correction was performed using Bonferroni method (*: adjust p<0.05).

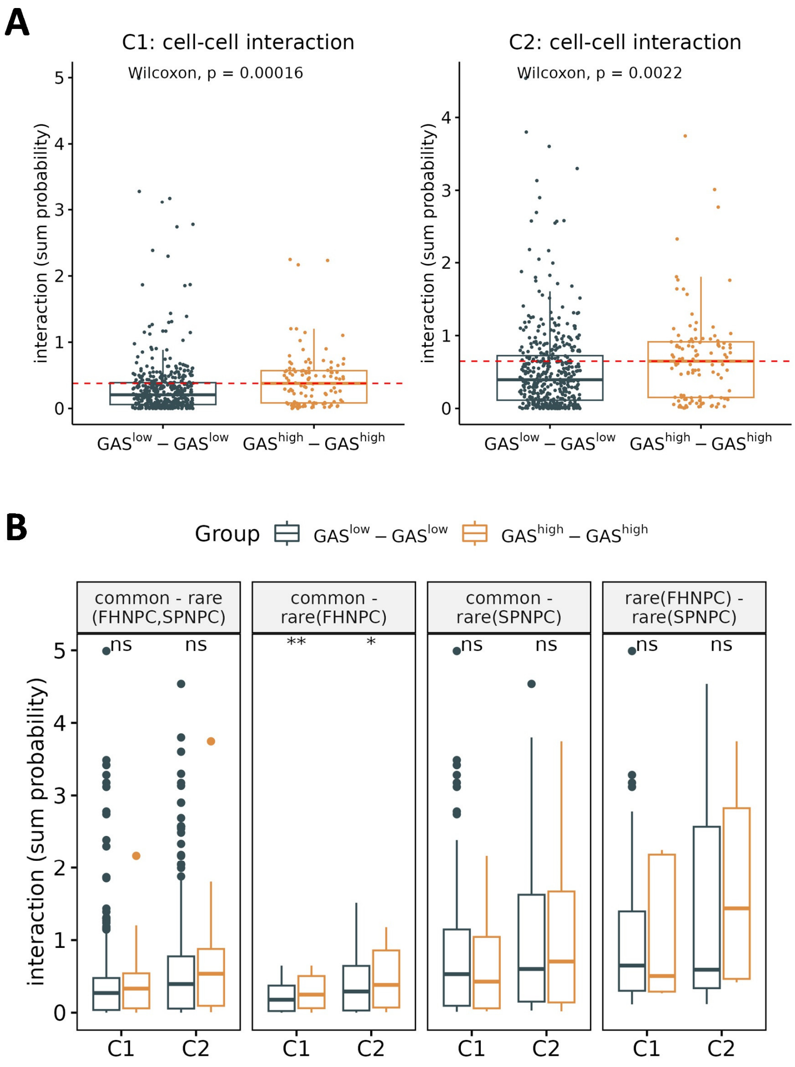


**Figure S6. Difference of interaction strength between $GAS^{high}$-$GAS^{high}$ and $GAS^{low}$-$GAS^{low}$ groups.** C1: cohort 1, C2: cohort 2. **A.** The results of the Wilcoxon tests comparing the cell-cell interaction strength (measured as sum probability) between $GAS^{high}$-$GAS^{high}$ and $GAS^{low}$-$GAS^{low}$ sub cell type pairs in two scRNA-seq cohorts (C1 and C2). **B.** The results of the Wilcoxon tests specifically comparing the cell-cell interaction strength (measured as sum probability) amongst cell subtype pair categories defined based on GASs for common and rare variant risk in C1 or C2 cohorts. 'common-rare(FHNPC,SPNPC)': the comparison of summed interaction probability between '$GAS^{high(common\ variant)}$-$GAS^{high(rare\ variant)}$' cell type pairs and '$GAS^{low(common\ variant)}$-$GAS^{low(rare\ variant)}$)' cell type pairs. '$GAS^{high(common\ variant)}$-$GAS^{high(rare\ variant)}$': the cell type pairs in which cell type 1 is rated as $GAS^{high}$ for common variant risk and cell type 2 is rated as $GAS^{high}$ for rare variant risk for NPC (FHNPC- or SPNPC- associated). '$GAS^{low(common\ variant)}$-$GAS^{low(rare\ variant)}$)': the cell type pairs in which cell type 1 is rated as $GAS^{low}$ for common variant risk and cell type 2 is rated as $GAS^{low}$ for rare variant risk for NPC (FHNPC- or SPNPC- associated). The same principles were applied to 'rare-rare', 'common-rare (FHNPC)', 'common-rare(SPNPC)' comparisons. 'rare (FHNPC)': the GASs for rare variant risk were calculated using data for NPC cases with family history. 'rare (SPNPC)': the GASs for rare variant risk were calculated using data for sporadic NPC cases.

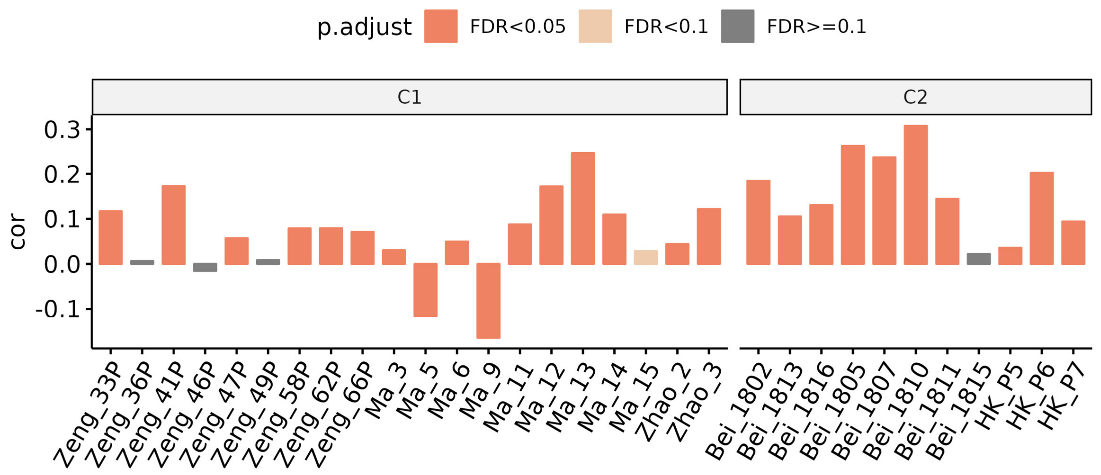


**Figure S7. Sample-specific correlation between the level of genetic association (measured as summed mean GASs) and the strength of cell-cell interaction (measured as sum probability).** Correlation (Spearman method) between the level of genetic association (measured as summed mean GASs) and cell-cell interaction levels (measured as sum probability) of all sub cell type pairs in each scRNA-seq sample (N=31). Multiple testing correction was performed using FDR method. C1: cohort 1, C2: cohort 2.

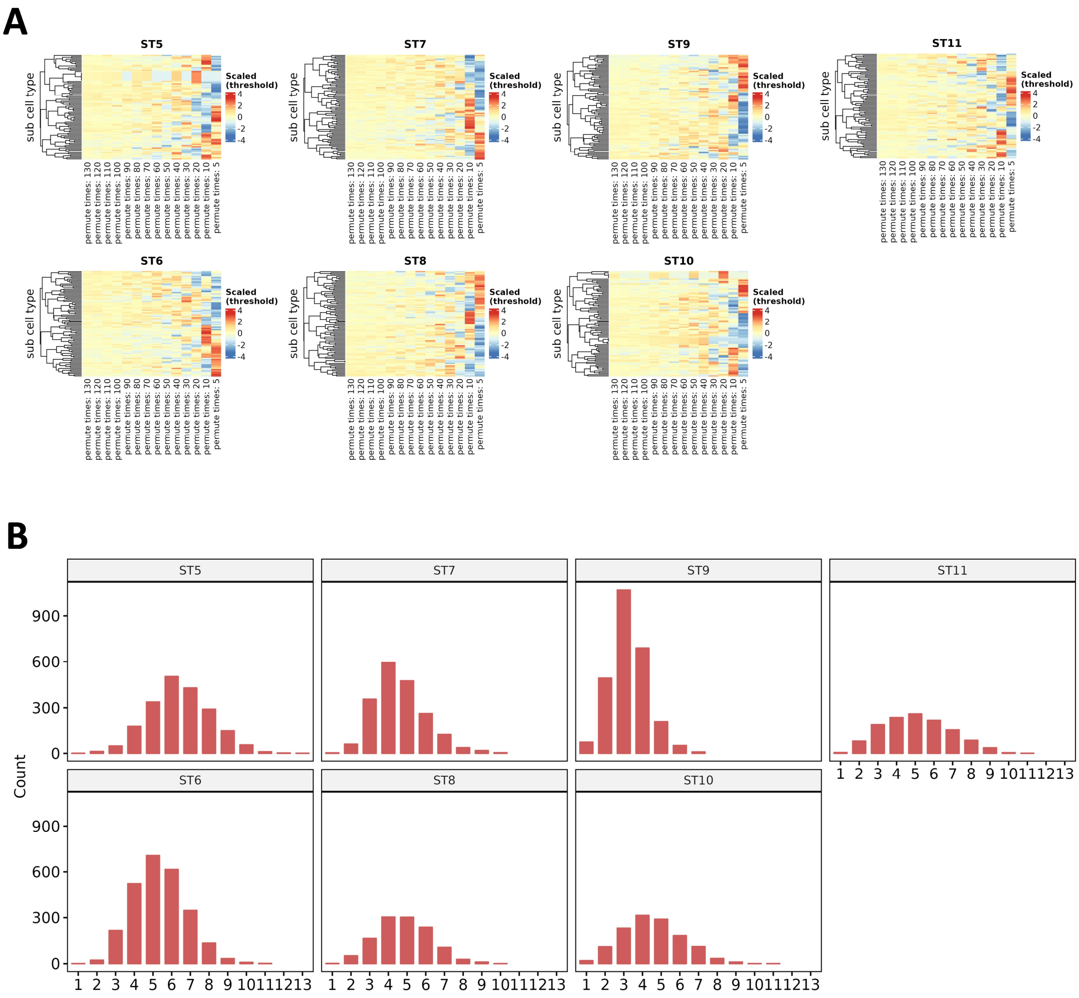


**Figure S8. Metrics of cell type identification for spots in spatial transcriptome samples. A.** Heatmaps showed the scaled threshold of cell type existence for 85 sub cell types identified by different permutation number setting (5, 10, 20, 30, 40, 50, 60, 70, 80, 90, 100, 110, 120 and 130) for the RCTD analysis in 7 spatial transcriptome samples. Scaling is performed on the row level (centered to 0). **B.** Barplots showed the distribution of the number of annotated sub cell types per spot in each spatial transcriptome sample.

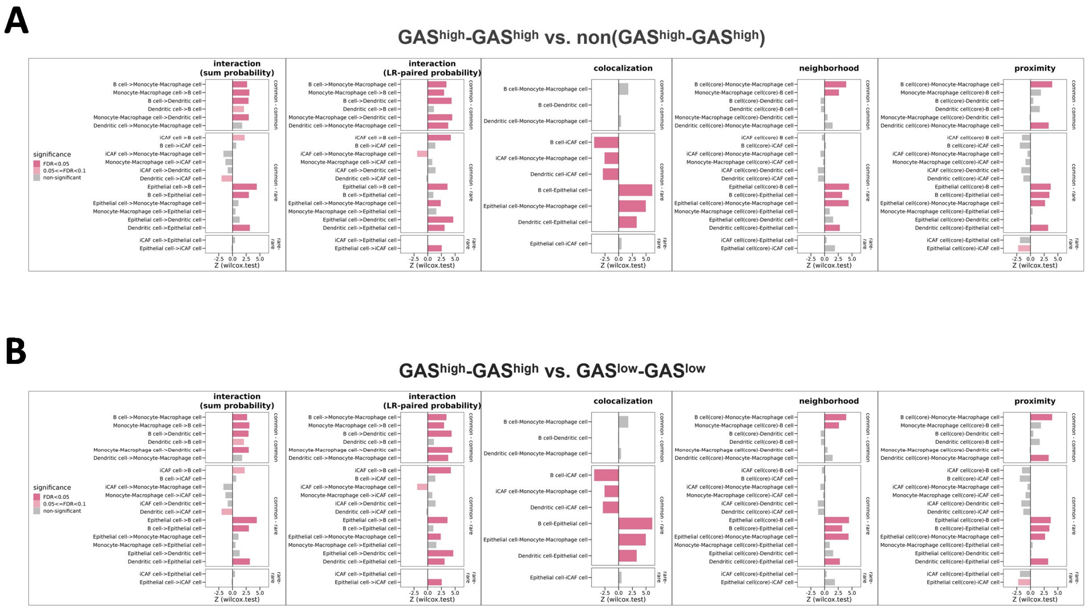


**Figure S9. Differentiated interaction strength and spatial distance between genetically-stratified cell type pairs in matched major cell types.** Z (wilcox.test): The test statistic reflecting inter-group differences which derived from Wilcoxon test. Multiple testing correction was performed using FDR method. The interaction strength was measured as sum probability and LR-paired probability, the spatial distance was measured as colocalization, neighborhood and proximity scores. **A.** The comparison was between $GAS^{high}$-$GAS^{high}$ and non($GAS^{high}$-$GAS^{high}$) groups. **B.** The comparison was between $GAS^{high}$-$GAS^{high}$ and $GAS^{low}$-$GAS^{low}$ groups.

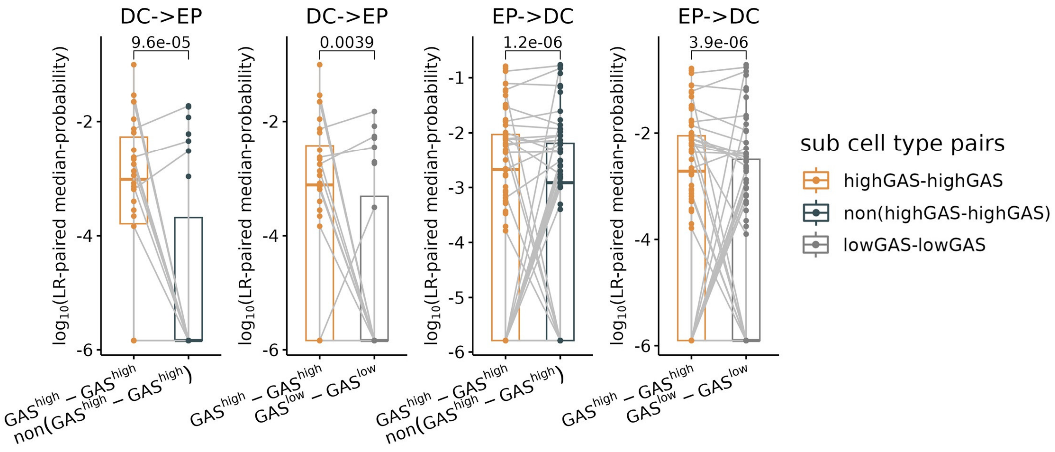


**Figure S10. Comparing LR-paired interaction between genetically-stratified dendritic-epithelial cell pairs.** Box plots compared LR-paired cell-cell interaction across different dendritic-epithelial cell pairs grouped into $GAS^{high}$-$GAS^{high}$, non($GAS^{high}$-$GAS^{high}$), $GAS^{low}$-$GAS^{low}$. Paired samples Wilcoxon tests were used to assess statistical differences.

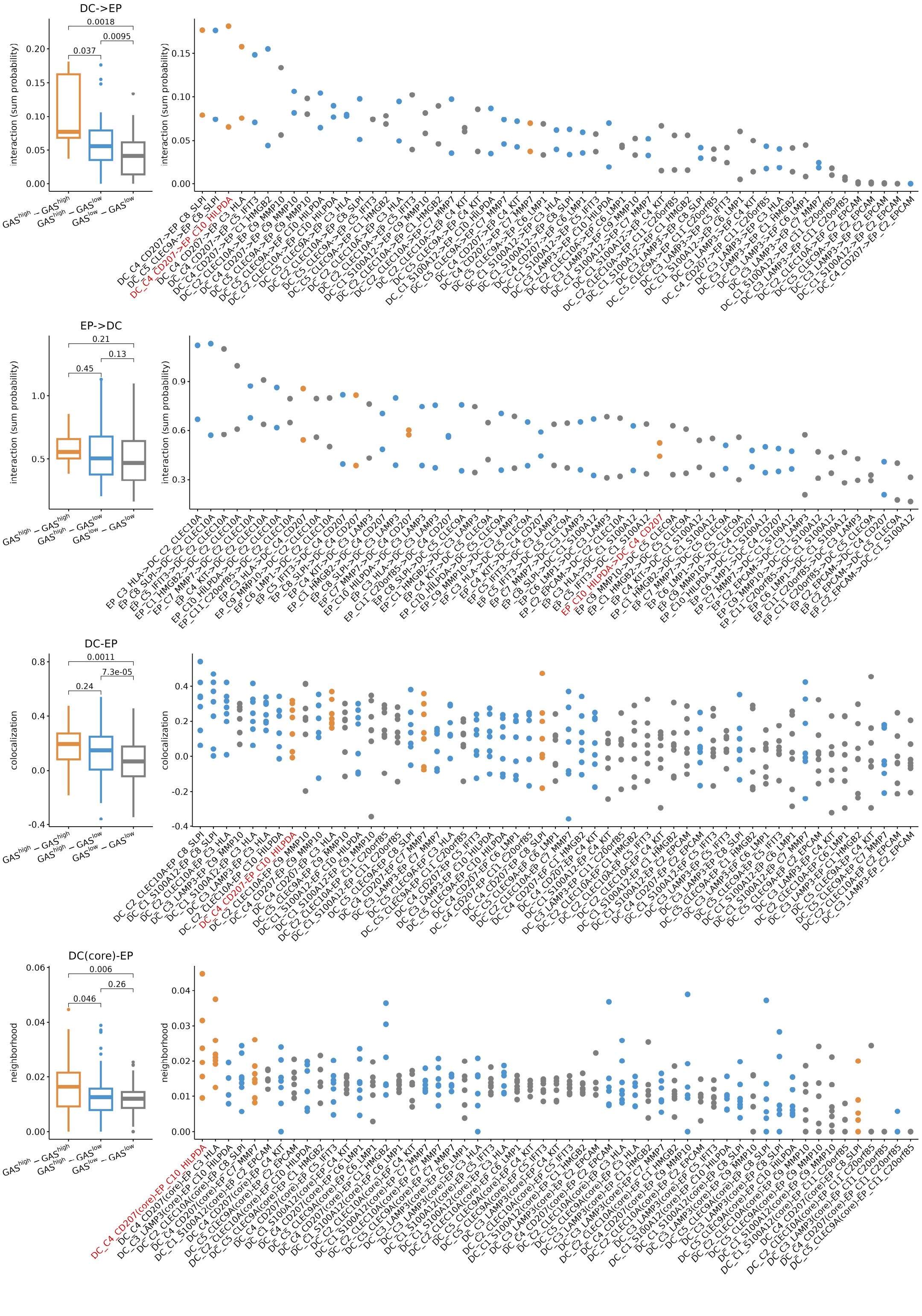
DC->EP
0.0018
0.0095
0.037
interaction (sum probability)
EP->DC
0.21
0.13
0.45
interaction (sum probability)
DC-EP
0.0011
7.3e-05
0.24
colocalization
DC(core)-EP
0.006
0.26
0.046
neighborhood
GAS^high − GAS^high
GAS^high − GAS^low
GAS^low − GAS^low

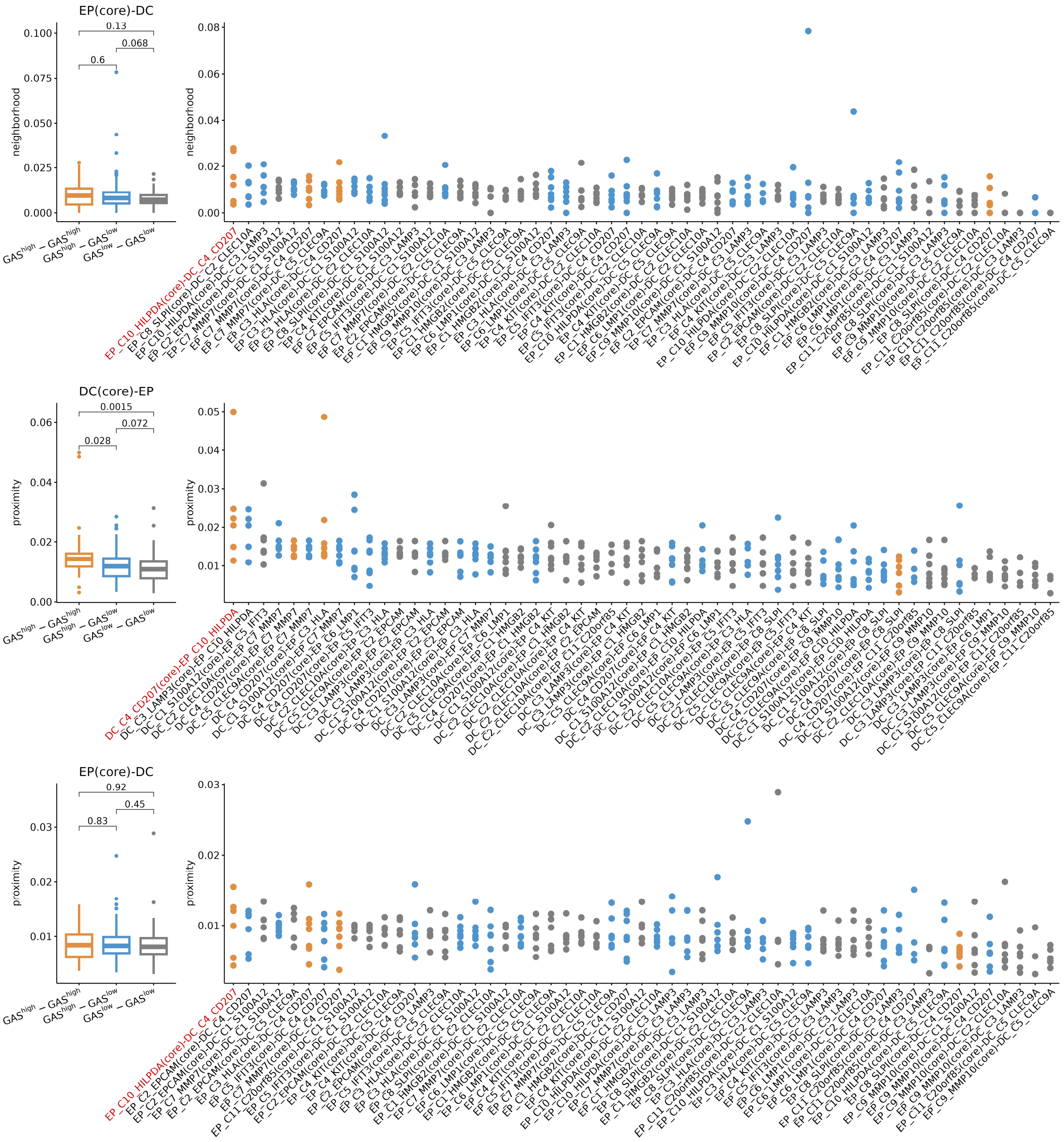


**Figure S11. Ranking dendritic and epithelial cell type pairs based on interaction and spatial distance metrics.** DC(core): represents the calculation of spatial distance metrics centered around DC sub cell type. EP(core): Represents the calculation of spatial distance metrics centered around EP sub cell type. DC->EP: Represents cell-cell interactions where the DC sub cell type expresses ligands, and the EP sub cell type expresses receptors. EP->DC: Represents cell-cell interactions where the EP sub cell type expresses ligands, and the DC sub cell type expresses receptors. The results of Wilcoxon test comparing cell-cell interaction strength (sum probability) and spatial distances (including "colocaliztion", "neighborhood" and "proximity" scores) between $GAS^{high}$-$GAS^{high}$, $GAS^{high}$-$GAS^{low}$, $GAS^{low}$-$GAS^{low}$ sub cell type pairs were also shown. The DC_C4_CD207-EP_C10_HILPDA pair was marked in red.

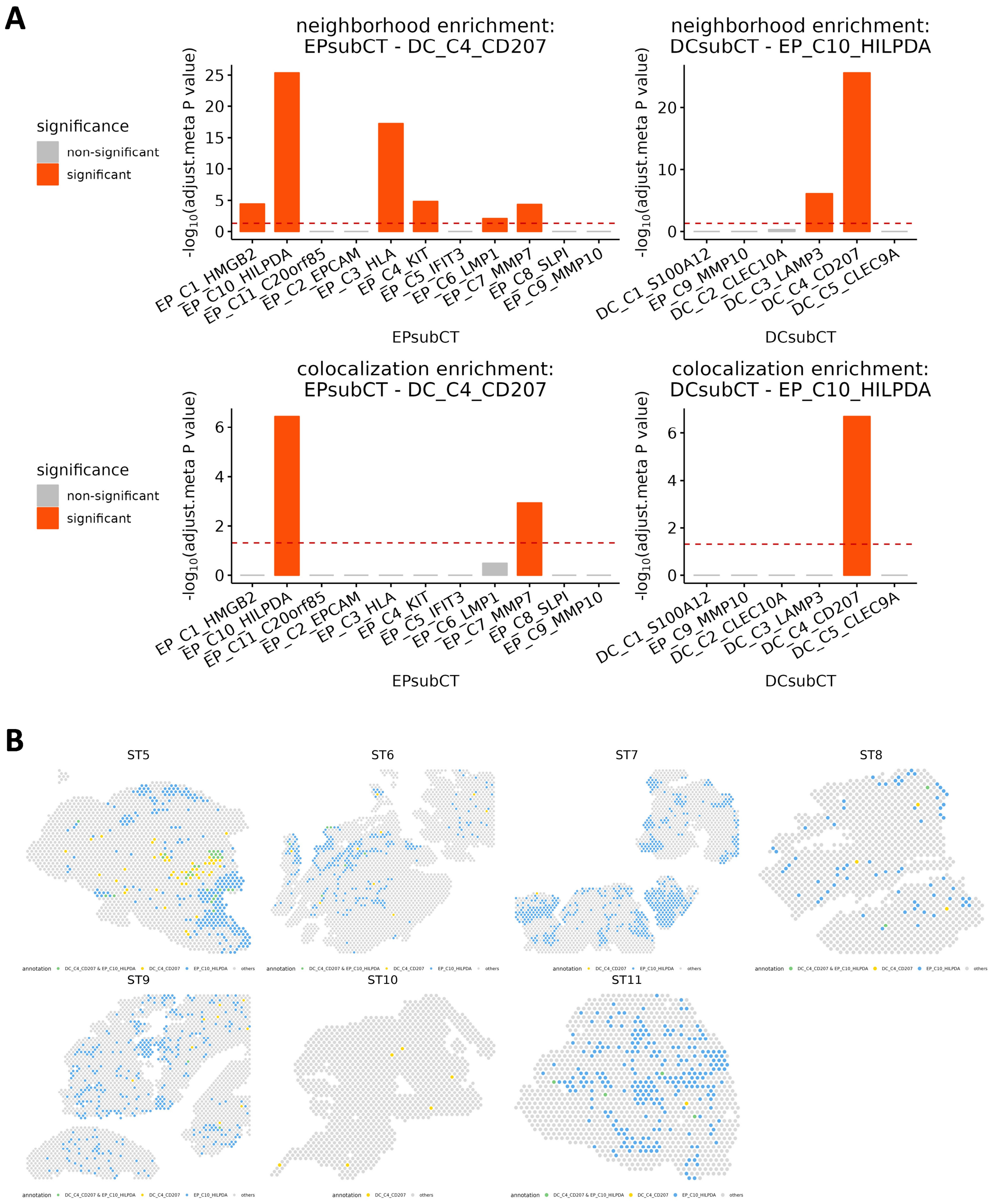


**Figure S12. Statistical tests and visualization of the colocalization and neighborhood relationship between DC_C4_CD207 and EP_C10_HILPDA. A.** The results of meta-analyzed Fisher's exact tests for the significant colocalization and neighborhood relationship between DC_C4_CD207 and EP_C10_HILPDA, and other sub cell types in dendritic and epithelial cell groups across samples (N=7) (see "Significant tests for the spatial association between two cell types" section in Method)**.** EPsubCT: Sub cell type of epithelial cells (EP); DCsubCT: Sub cell type of dendritic cell (DC). Multiple testing correction was performed using Bonferroni method. **B.** Visualization of the spots in spatial transcriptome data of the 7 samples.

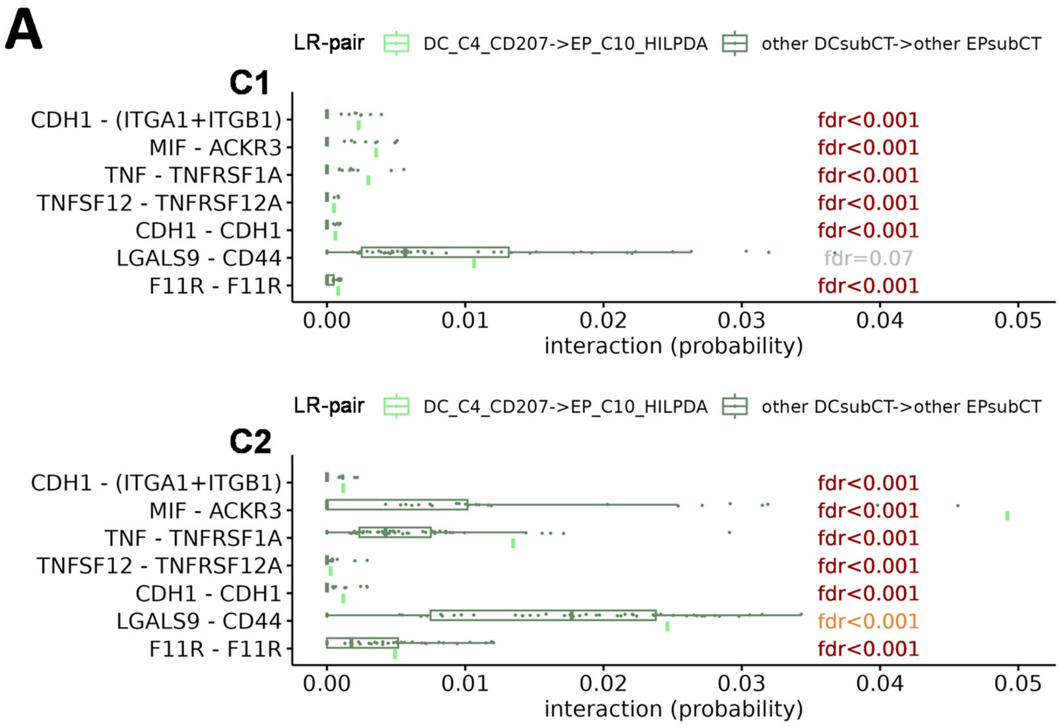
A
LR-pair
DC_C4_CD207->EP_C10_HILPDA
other DCsubCT->other EPsubCT
C1
CDH1 - (ITGA1+ITGB1)
MIF - ACKR3
TNF - TNFRSF1A
TNFSF12 - TNFRSF12A
CDH1 - CDH1
LGALS9 - CD44
F11R - F11R
fdr<0.001
fdr<0.001
fdr<0.001
fdr<0.001
fdr<0.001
fdr=0.07
fdr<0.001
0.00
0.01
0.02
0.03
0.04
0.05
interaction (probability)
LR-pair
DC_C4_CD207->EP_C10_HILPDA
other DCsubCT->other EPsubCT
C2
CDH1 - (ITGA1+ITGB1)
MIF - ACKR3
TNF - TNFRSF1A
TNFSF12 - TNFRSF12A
CDH1 - CDH1
LGALS9 - CD44
F11R - F11R
fdr<0.001
fdr<0.001
fdr<0.001
fdr<0.001
fdr<0.001
fdr<0.001
fdr<0.001
0.00
0.01
0.02
0.03
0.04
0.05
interaction (probability)

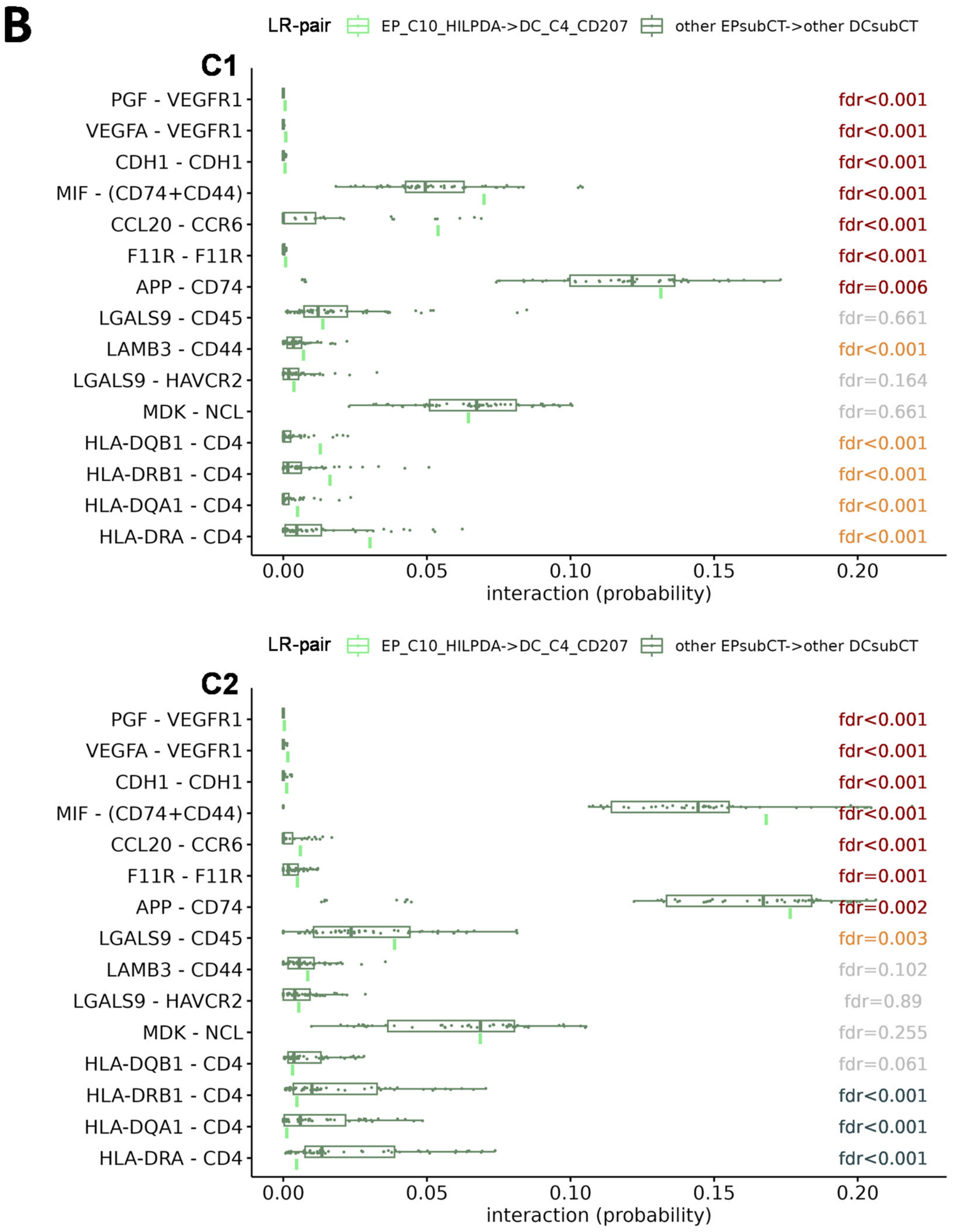


**Figure S13. Examining the significantly interacted ligand(L)-receptor(R) pairs in target cell type-pair (DC_C4_CD207-EP_C10_HILPDA) and other cell type pairs within matched major cell types (dendritic and epithelial cells).** C1: cohort1, C2: cohort 2. Corrected P-value (FDR) of red or orange note representing significantly increased interaction probability ($P_{FDR}$<0.05) in the DC_C4_CD207-EP_C10_HILPDA cell type pair compared to others in two cohorts (C1 and C2) or one cohort, respectively. A total of 11 unique LR pairs with such increased interaction probability were identified as follows: CDH1-(ITGA1+ITGB1), MIF-ACKR3, TNF-TNFRSF1A, TNFSF12-TNFRSF12A, PGF-VEGFR1, VEGFA-VEGFR1, MIF-(CD74+CD44), CCL20-CCR6, APP-CD74, CDH1-CDH1, F11R-F11R. Corrected P-value (FDR) of blue-black note representing significantly decreased interaction probability ($P_{FDR}$<0.05) in the DC_C4_CD207-EP_C10_HILPDA cell type pair compared to others in the cohort. Corrected P-value (FDR) of grey note representing no significant different between the interaction probability of DC_C4_CD207-EP_C10_HILPDA cell type pair and others in the cohort. **A.** Significantly interacted LR-pairs where the ligand gene is expressed in DC_C4_CD207 and the receptor gene is expressed in EP_C10_HILPDA. **B.** Significantly interacted LR-pairs where the receptor gene is expressed in DC_C4_CD207 and the ligand gene is expressed in EP_C10_HILPDA.

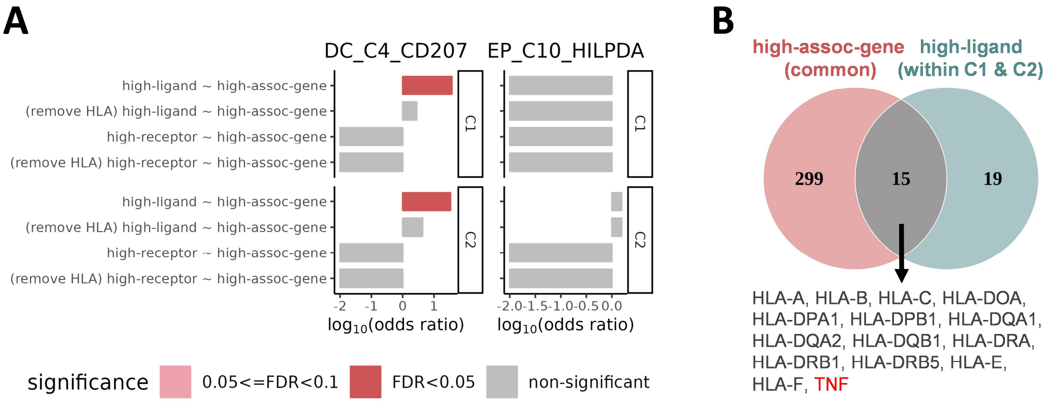


**Figure S14. Test for direct contribution of genetic associations to the cell-cell interaction between DC_C4_CD207 and EP_C10_HILPDA cell types under alternative threshold for significance of genetic association and interaction (P<0.005).** C1: cohort 1, C2: cohort 2. **A.** Direct participation: Fisher's exact test results examining the significance of the overlap between genetically associated genes (P<0.005 in original GWAS or EWAS. Labeled as “high-assoc-gene”) and ligand (L)/receptor (R) genes of significantly interacted LR-pairs (P<0.005. Labeled as “high-ligand”/ “high-receptor”) in DC_C4_CD207 or EP_C10_HILPDA cell type. Multiple testing correction for Fisher's exact test results was performed using FDR method. **B.** Intersection of NPC-associated genes derived from common variants (P<0.005) and ligand genes of significantly interacted LR-pairs (P<0.005) in DC_C4_CD207. The gene marked in red is a ligand gene of the key LR- pairs contributing to the strong crosstalk between DC_C4_CD207 and EP_C10_HILPDA crosstalk. common: the NPC-associated susceptibility genes derived from common variants; within C1 & C2: the ligand genes of significantly interacting LR pairs in both C1 and C2 scRNA-seq cohorts.

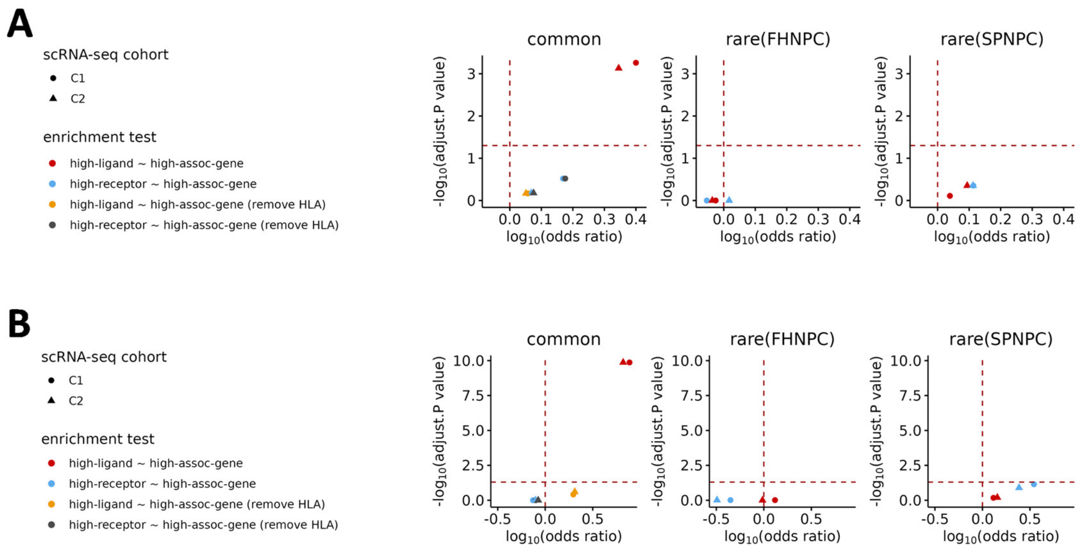


**Figure S15. Enrichment test results for the overlap between NPC-associated genes (label as 'high-assoc-gene') and ligand (L) or receptor genes of significant interacted LR-pairs using different significance thresholds (P<0.05 or P<0.005) for genetic association and interaction.** C1: cohort 1, C2: cohort 2. P-values and odds ratios were estimated from Fisher's exact test. Multiple testing correction for Fisher's exact test results was performed using FDR method. Common, rare(FHNPC), rare(SPNPC): the genetic association statistics was derived based on common, rare(FHNPC), rare(SPNPC) variants, respectively. The included ligand/receptor (high-ligand/high-receptor) pairs were those significantly interacting with others in at least one cell type. **A.** Fisher's exact test results examining the significance of the overlap between NPC-associated genes (P<0.05 in original GWAS or EWAS; labeled as "high-assoc-gene") and ligand (L)/receptor (R) genes encoding significantly interacted LR-pairs (P<0.05; labeled as "high-ligand"/ "high-receptor"). **B.** Fisher's exact test results examining the significance of the overlap between NPC-associated genes (P<0.005 in original GWAS or EWAS; labeled as "high-assoc-gene") and ligand (L)/receptor (R) genes encoding significantly interacted LR-pairs (P<0.005; labeled as "high-ligand"/ "high-receptor").

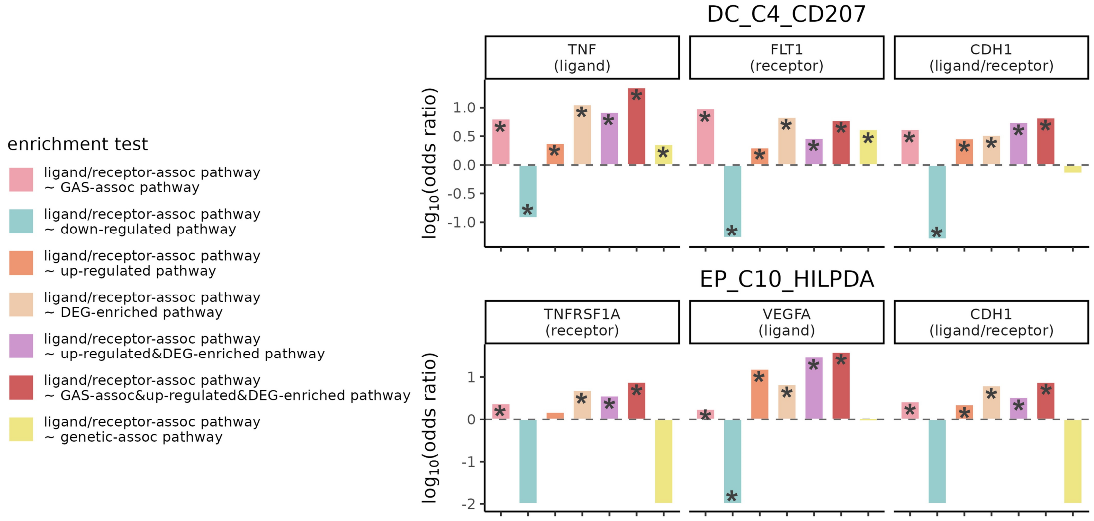


**Figure S16. Enrichment test for the gene overlap between multiple pathways involving ligand/receptor interactions, genetic signals, or featured functions of specific sub cell types.** Fisher's exact test results examining the significance of the pathway overlap between LR-associated pathways (ligand/receptor-assoc pathways) with NPC genetically associated pathways (genetic-assoc pathways), with activity up-/down-regulated pathways, with DEG-enriched pathways and with GAS-assoc pathways (Details in Method about the definition of the above pathways) in DC_C4_CD207 or EP_C10_HILPDA cell type. *: adjust p<0.05. Multiple testing correction was performed using FDR method.

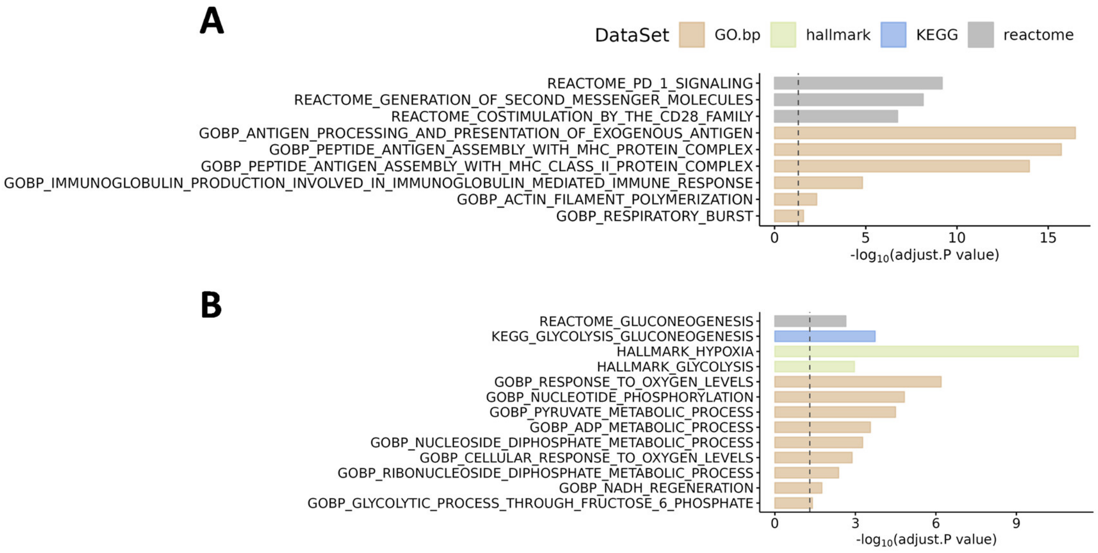


**Figure S17. Pathways significantly enriched in differentially expressed genes (DEGs) and upregulated in DC_C4_CD207 or EP_C10_HILPDA cell types. A.** Pathways specifically upregulated in DC_C4_CD207 compared to other DC sub cell types, and significantly overrepresented in DEGs of DC_C4_CD207. **B.** Pathways specifically upregulated in EP_C10_HILPDA compared to other EP sub cell types, and significantly overrepresented in DEGs of EP_C10_HILPDA. Pathways from GO biological process, hallmark, KEGG and Reactome databases in MSigDB were displayed. Multiple testing correction for each database was performed using Bonferroni method.

A

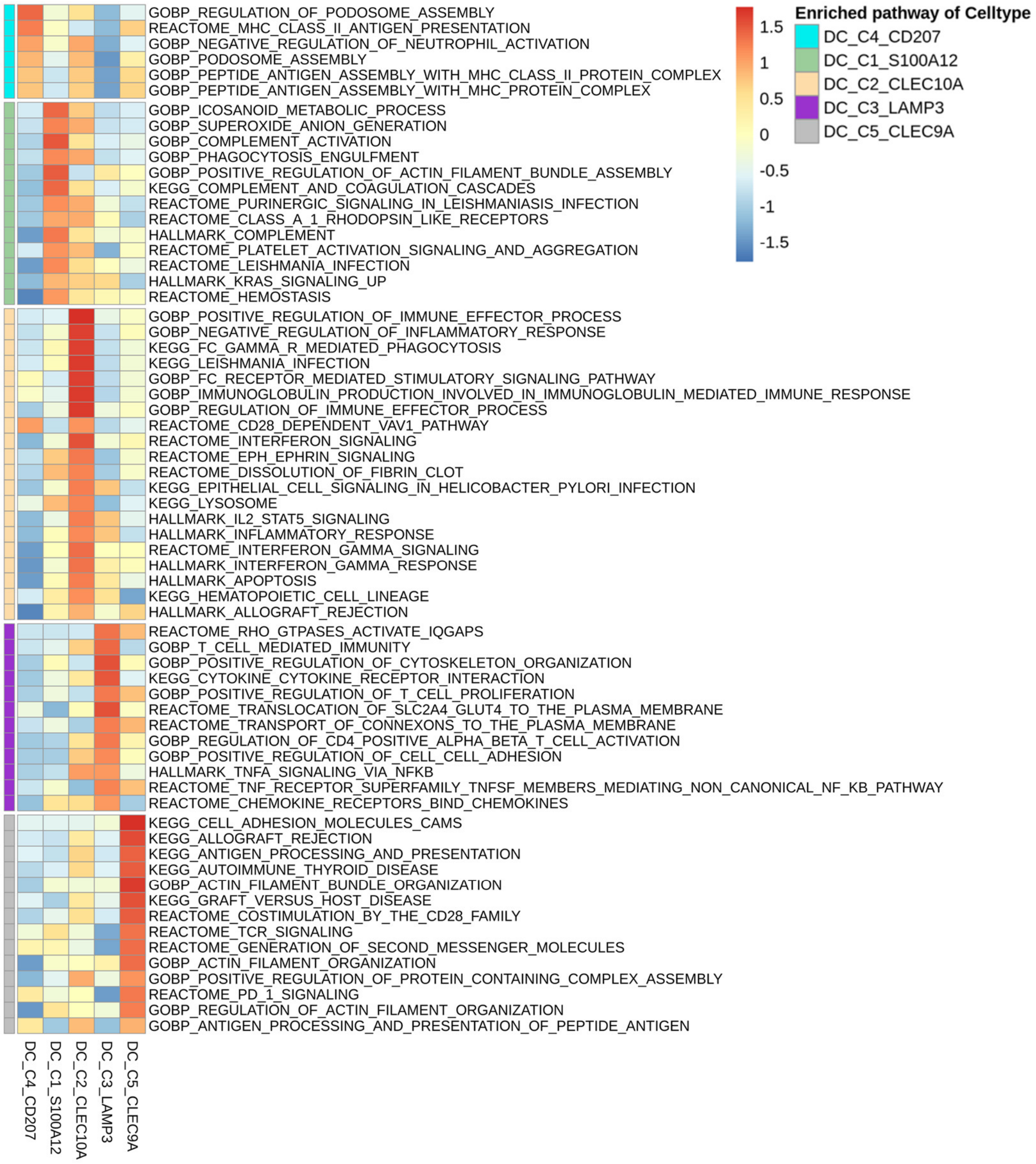

GOBP_REGULATION_OF_PODOSOME_ASSEMBLY
REACTOME_MHC_CLASS_II_ANTIGEN_PRESENTATION
GOBP_NEGATIVE_REGULATION_OF_NEUTROPHIL_ACTIVATION
GOBP_PODOSOME_ASSEMBLY
GOBP_PEPTIDE_ANTIGEN_ASSEMBLY_WITH_MHC_CLASS_II_PROTEIN_COMPLEX
GOBP_PEPTIDE_ANTIGEN_ASSEMBLY_WITH_MHC_PROTEIN_COMPLEX
GOBP_ICOSANOID_METABOLIC_PROCESS
GOBP_SUPEROXIDE_ANION_GENERATION
GOBP_COMPLEMENT_ACTIVATION
GOBP_PHAGOCYTOSIS_ENGULFMENT
GOBP_POSITIVE_REGULATION_OF_ACTIN_FILAMENT_BUNDLE_ASSEMBLY
KEGG_COMPLEMENT_AND_COAGULATION_CASCADES
REACTOME_PURINERGIC_SIGNALING_IN_LEISHMANIASIS_INFECTION
REACTOME_CLASS_A_1_RHODOPSIN_LIKE_RECEPTORS
HALLMARK_COMPLEMENT
REACTOME_PLATELET_ACTIVATION_SIGNALING_AND_AGGREGATION
REACTOME_LEISHMANIA_INFECTION
HALLMARK_KRAS_SIGNALING_UP
REACTOME_HEMOSTASIS
GOBP_POSITIVE_REGULATION_OF_IMMUNE_EFFECTOR_PROCESS
GOBP_NEGATIVE_REGULATION_OF_INFLAMMATORY_RESPONSE
KEGG_FC_GAMMA_R_MEDIATED_PHAGOCYTOSIS
KEGG_LEISHMANIA_INFECTION
GOBP_FC_RECEPTOR_MEDIATED_STIMULATORY_SIGNALING_PATHWAY
GOBP_IMMUNOGLOBULIN_PRODUCTION_INVOLVED_IN_IMMUNOGLOBULIN_MEDIATED_IMMUNE_RESPONSE
GOBP_REGULATION_OF_IMMUNE_EFFECTOR_PROCESS
REACTOME_CD28_DEPENDENT_VAV1_PATHWAY
REACTOME_INTERFERON_SIGNALING
REACTOME_EPH_EPHRIN_SIGNALING
REACTOME_DISSOLUTION_OF_FIBRIN_CLOT
KEGG_EPITHELIAL_CELL_SIGNALING_IN_HELICOBACTER_PYLORI_INFECTION
KEGG_LYSOSOME
HALLMARK_IL2_STAT5_SIGNALING
HALLMARK_INFLAMMATORY_RESPONSE
REACTOME_INTERFERON_GAMMA_SIGNALING
HALLMARK_INTERFERON_GAMMA_RESPONSE
HALLMARK_APOPTOSIS
KEGG_HEMATOPOIETIC_CELL_LINEAGE
HALLMARK_ALLOGRAFT_REJECTION
REACTOME_RHO_GTPASES_ACTIVATE_IQGAPS
GOBP_T_CELL_MEDIATED_IMMUNITY
GOBP_POSITIVE_REGULATION_OF_CYTOSKELETON_ORGANIZATION
KEGG_CYTOKINE_CYTOKINE_RECEPTOR_INTERACTION
GOBP_POSITIVE_REGULATION_OF_T_CELL_PROLIFERATION
REACTOME_TRANSLOCATION_OF_SLC2A4_GLUT4_TO_THE_PLASMA_MEMBRANE
REACTOME_TRANSPORT_OF_CONNEXONS_TO_THE_PLASMA_MEMBRANE
GOBP_REGULATION_OF_CD4_POSITIVE_ALPHA_BETA_T_CELL_ACTIVATION
GOBP_POSITIVE_REGULATION_OF_CELL_CELL_ADHESION
HALLMARK_TNFA_SIGNALING_VIA_NFKB
REACTOME_TNF_RECEPTOR_SUPERFAMILY_TNFSF_MEMBERS_MEDIATING_NON_CANONICAL_NF_KB_PATHWAY
REACTOME_CHEMOKINE_RECEPTORS_BIND_CHEMOKINES
KEGG_CELL_ADHESION_MOLECULES_CAMS
KEGG_ALLOGRAFT_REJECTION
KEGG_ANTIGEN_PROCESSING_AND_PRESENTATION
KEGG_AUTOIMMUNE_THYROID_DISEASE
GOBP_ACTIN_FILAMENT_BUNDLE_ORGANIZATION
KEGG_GRAFT_VERSUS_HOST_DISEASE
REACTOME_COSTIMULATION_BY_THE_CD28_FAMILY
REACTOME_TCR_SIGNALING
REACTOME_GENERATION_OF_SECOND_MESSENGER_MOLECULES
GOBP_ACTIN_FILAMENT_ORGANIZATION
GOBP_POSITIVE_REGULATION_OF_PROTEIN_CONTAINING_COMPLEX_ASSEMBLY
REACTOME_PD_1_SIGNALING
GOBP_REGULATION_OF_ACTIN_FILAMENT_ORGANIZATION
GOBP_ANTIGEN_PROCESSING_AND_PRESENTATION_OF_PEPTIDE_ANTIGEN
DC_C4_CD207
DC_C1_S100A12
DC_C2_CLEC10A
DC_C3_LAMP3
DC_C5_CLEC9A
1.5
1
0.5
0
-0.5
-1
-1.5
Enriched pathway of Celltype
DC_C4_CD207
DC_C1_S100A12
DC_C2_CLEC10A
DC_C3_LAMP3
DC_C5_CLEC9A

**B**

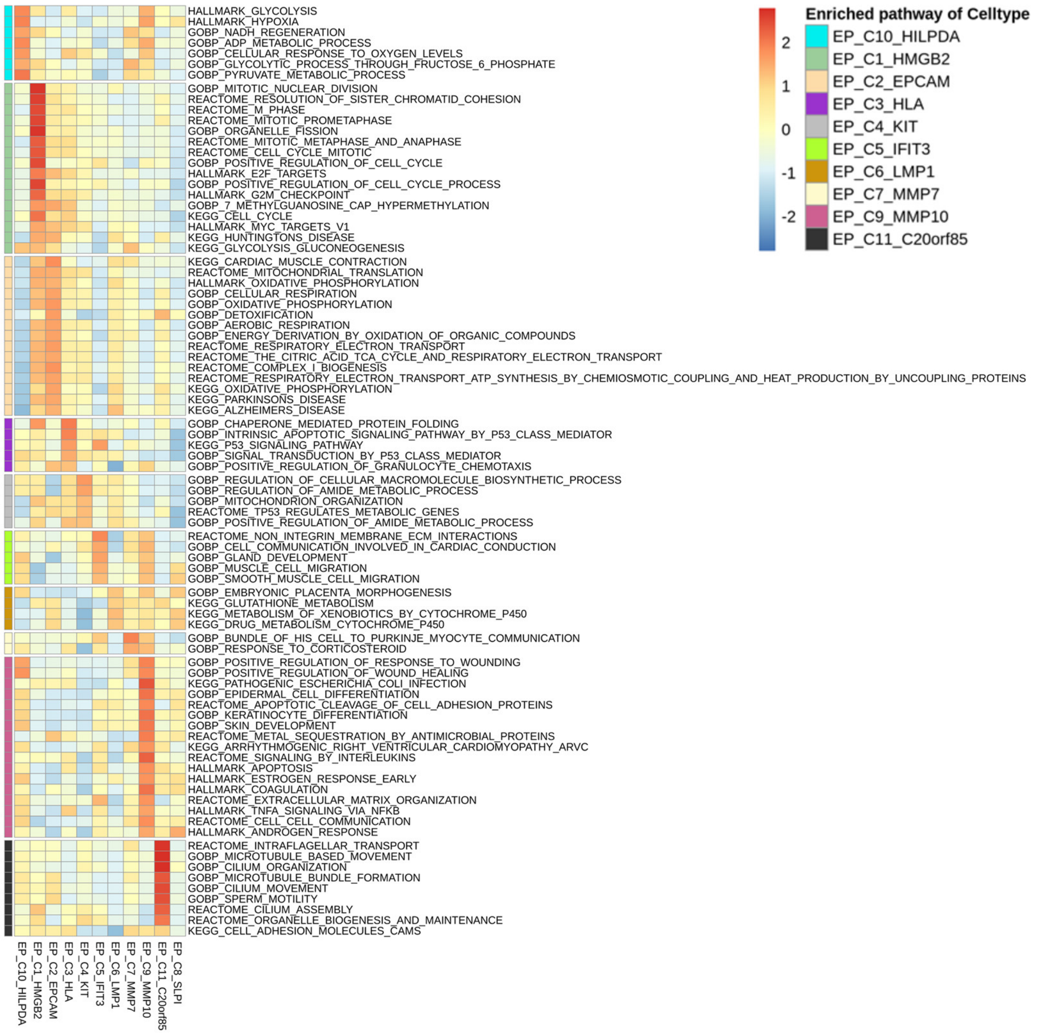


**Figure S18. Functional characterization and comparison of sub cell types within Dendritic cells (DC) and Epithelial cells (EP). A.** Heatmap of GSVA scores (scaled by row) showing the up-regulated signaling pathways of Dendritic cells (DC) sub cell types. **B.** Heatmap of GSVA scores (scaled by row) showing the up-regulated signaling pathways of Epithelial cells (EP) sub cell types. The group "Enriched pathway of Celltype" means the marked pathway was significantly overrepresented in differentially expressed genes (DEGs) of the annotated sub cell type. Pathways from GO biological process, hallmark, KEGG and Reactome databases in MSigDB were used, and for each sub cell type, only top 5 up-regulated pathways of each database were displayed.

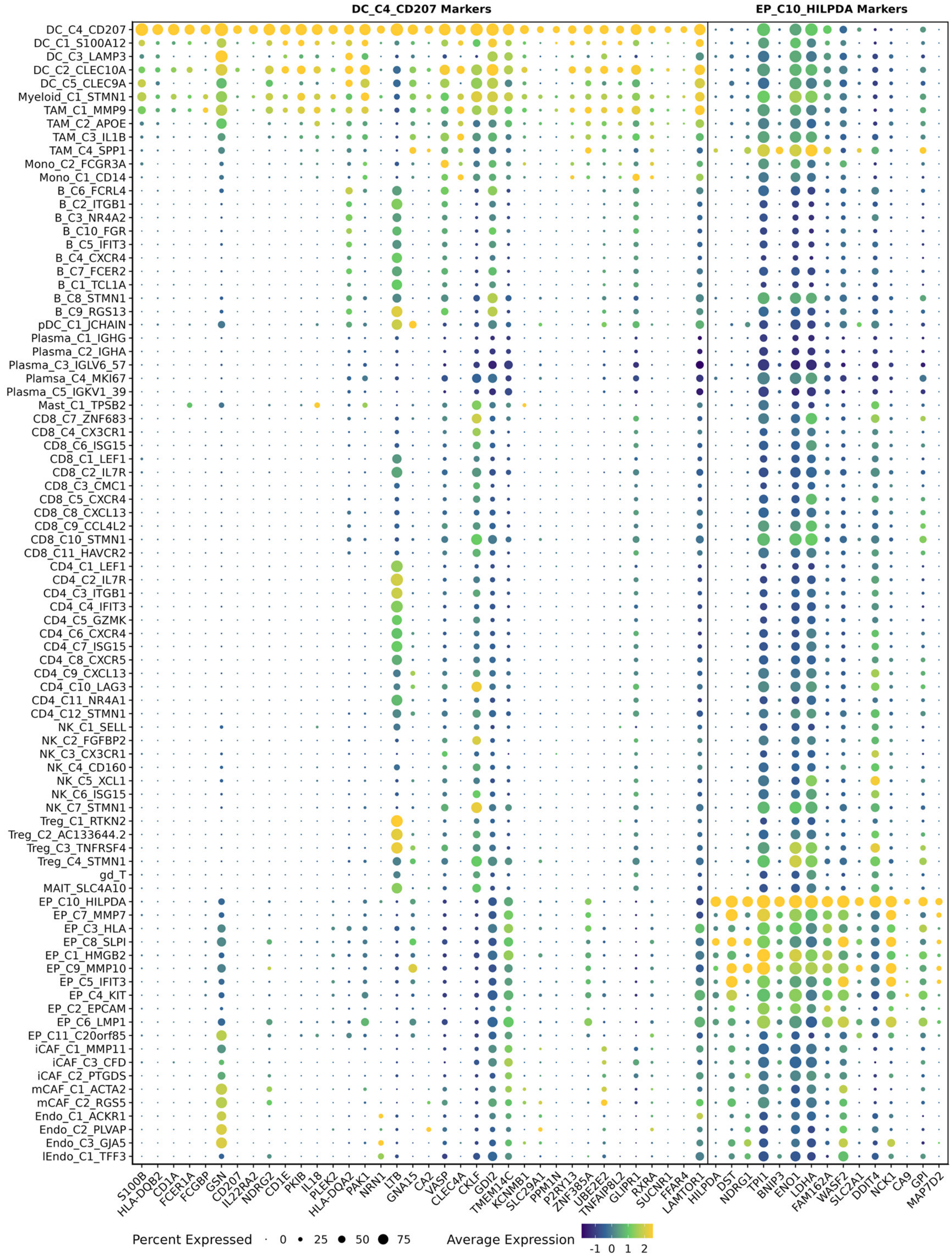


**Figure S19. Expression of marker genes in DC_C4_CD207 and EP_C10_HILPDA.** The plot shows the percentage of cells expressing and the average expression level of DC_C4_CD207 (left) and EP_C10_HILPDA (right) marker genes across all sub cell types. These genes were selected as markers and were used to estimate the infiltration levels of DC_C4_CD207 and EP_C10_HILPDA in bulk RNA-seq data for subsequent survival analysis.

A

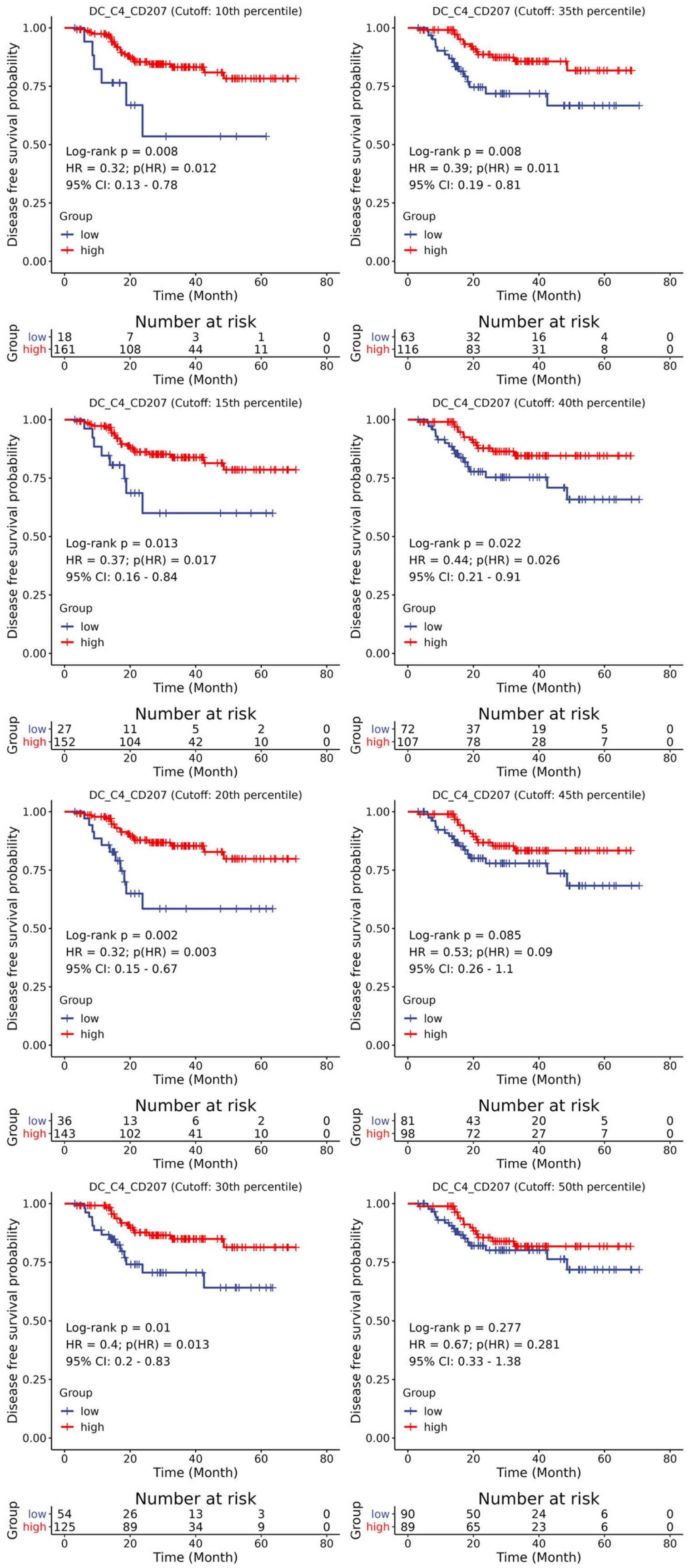
DC_C4_CD207 (Cutoff: 10th percentile)
Log-rank p = 0.008
HR = 0.32; p(HR) = 0.012
95% CI: 0.13 - 0.78
Number at risk
low 18 7 3 1 0
high 161 108 44 11 0
DC_C4_CD207 (Cutoff: 35th percentile)
Log-rank p = 0.008
HR = 0.39; p(HR) = 0.011
95% CI: 0.19 - 0.81
Number at risk
low 63 32 16 4 0
high 116 83 31 8 0
DC_C4_CD207 (Cutoff: 15th percentile)
Log-rank p = 0.013
HR = 0.37; p(HR) = 0.017
95% CI: 0.16 - 0.84
Number at risk
low 27 11 5 2 0
high 152 104 42 10 0
DC_C4_CD207 (Cutoff: 40th percentile)
Log-rank p = 0.022
HR = 0.44; p(HR) = 0.026
95% CI: 0.21 - 0.91
Number at risk
low 72 37 19 5 0
high 107 78 28 7 0
DC_C4_CD207 (Cutoff: 20th percentile)
Log-rank p = 0.002
HR = 0.32; p(HR) = 0.003
95% CI: 0.15 - 0.67
Number at risk
low 36 13 6 2 0
high 143 102 41 10 0
DC_C4_CD207 (Cutoff: 45th percentile)
Log-rank p = 0.085
HR = 0.53; p(HR) = 0.09
95% CI: 0.26 - 1.1
Number at risk
low 81 43 20 5 0
high 98 72 27 7 0
DC_C4_CD207 (Cutoff: 30th percentile)
Log-rank p = 0.01
HR = 0.4; p(HR) = 0.013
95% CI: 0.2 - 0.83
Number at risk
low 54 26 13 3 0
high 125 89 34 9 0
DC_C4_CD207 (Cutoff: 50th percentile)
Log-rank p = 0.277
HR = 0.67; p(HR) = 0.281
95% CI: 0.33 - 1.38
Number at risk
low 90 50 24 6 0
high 89 65 23 6 0
Disease free survival probability
Time (Month)
Group
low
high

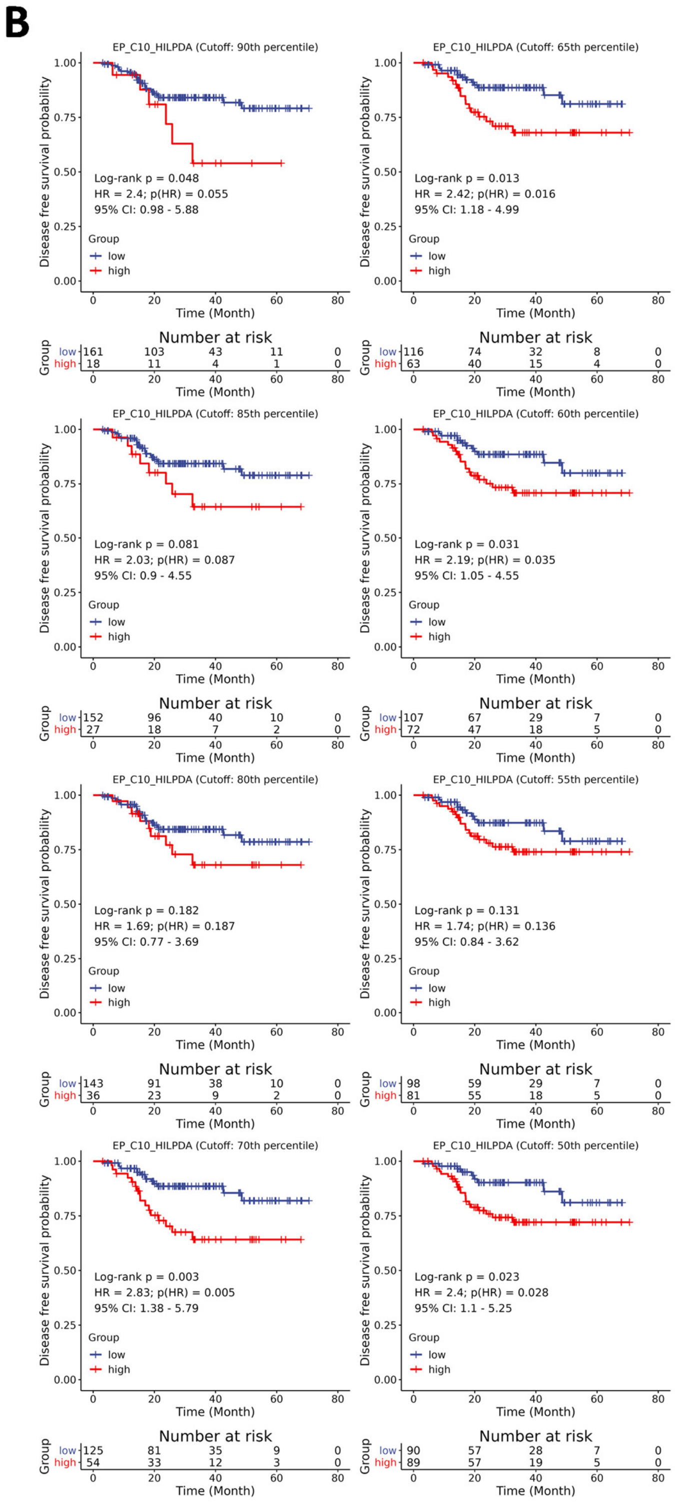

**Figure S20. Survival analyses for infiltration levels of DC_C4_CD207 or EP_C10_HILPDA by using different percentiles as cut point of patients. A.** Kaplan-Meier survival analyses results of DC_C4_CD207 infiltration levels in two bulk RNA-seq cohorts. The 10th, 15th, 20th, 30th, 35th, 40th, 45th, and 50th percentiles of DC_C4_CD207 infiltration levels were used as cut points. **B.** Kaplan-Meier survival analyses results of EP_C10_HILPDA infiltration levels in two cohorts. The 90th, 85th, 80th, 70th, 65th, 60th, 55th, and 50th percentiles of EP_C10_HILPDA infiltration levels were used as cut points.

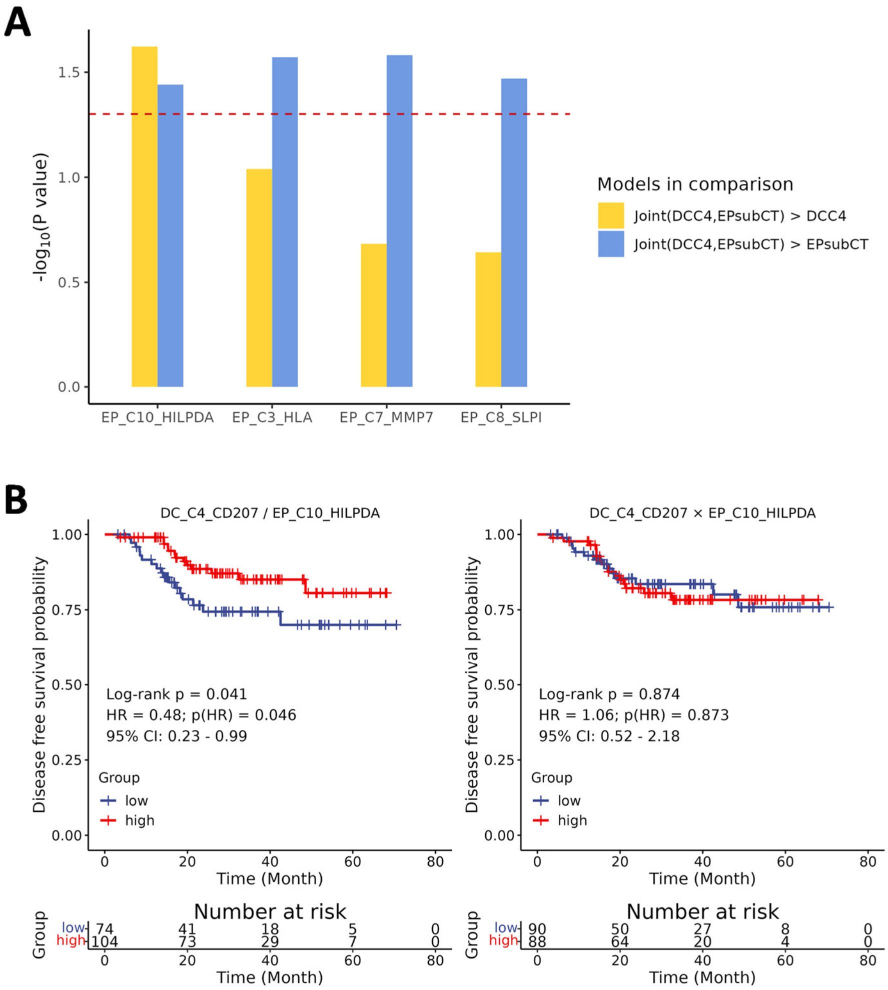


**Figure S21. Evaluation of the predictive performance for indexes/models predicting NPC prognosis by joint consideration of infiltration levels of DC_C4_CD207 and EP_C10_HILPDA. A.** Comparison of goodness-of-fit between Cox regression models with infiltration levels of DC_C4_CD207 or epithelial sub cell types accounted individually and jointly. using likelihood ratio test. The height of bars showed the significances of the likelihood ratio tests. DCC4: Cox regression model constructed based on the infiltration level of DC_C4_CD207. EPsubCT: Cox regression model constructed based on the infiltration level of a certain genetically-associated EP sub cell type. Joint(DCC4,EPsubCT): Cox regression model considering the infiltration levels of DC_C4_CD207 and a certain genetically-associated EP sub cell type. The P-value were derived from likelihood ratio test by comparing the goodness of fit between two models on both sides of ">" in the legend. The model on the right side of ">" is nested model of the model on the left. **B.** The left panel: Kaplan-Meier survival analysis result of an index calculated as DC_C4_CD207 infiltration levels divide by EP_C10_HILPDA infiltration levels. The right panel: Kaplan-Meier survival analysis result of an index calculated as DC_C4_CD207 infiltration levels multiply by EP_C10_HILPDA infiltration levels. For the ratio metric (DC_C4_CD207 / EP_C10_HILPDA), the cut-point was defined as dividing the 25th percentile of DC_C4_CD207 infiltration by the 75th percentile of EP_C10_HILPDA infiltration; for the product metric (DC_C4_CD207 × EP_C10_HILPDA), the cut-point was defined as multiplying the same percentile values.
